# Modelling the impact of long-acting monoclonal antibody, maternal vaccine and hybrid programs of RSV immunisation in temperate Western Australia

**DOI:** 10.64898/2026.03.02.26347477

**Authors:** Fiona Giannini, Alexandra B. Hogan, Christopher C Blyth, Kathryn Glass, Hannah C. Moore, the STAMP RSV investigator team

**Author notes:** Corresponding author: Fiona Giannini. joint senior authors (Kathryn Glass, Hannah C. Moore).

## Abstract

**Background:** Two RSV immunisations products: a maternal vaccine, Abrysvo, and a long-acting monoclonal antibody, nirsevimab, both designed to prevent RSV illness in infants, have recently become available. Modelling evidence is required to inform how to optimally use these products in immunisation programs to reduce the burden of RSV in young children.

**Methods:** We extend a dynamic transmission model calibrated to RSV-hospitalisation data of children aged < 5 years in temperate Western Australia (WA) to simulate a range of potential RSV immunisation programs. Using our model, we estimate the impact of both single-product and hybrid RSV immunisation programs. The analysis considers timing of administration, coverage levels and targeting of high-risk groups. Impact on RSV burden is analysed in the context of the WA setting and the possible significant cost differences between the two products.

**Results:** All programs analysed were effective in reducing RSV burden. Programs using nirsevimab for newborn infants at similar coverage levels to the Abrysvo programs, averted more RSV-hospitalisations annually. Seasonal programs that focused on protection during high RSV activity and programs targeting high-risk infants were the most efficient in reducing RSV burden. When dose cost is considered alongside program impact on RSV burden, we find evidence to support further economic analysis of hybrid programs as they could mitigate the cost differential between the two products while remaining highly effective in reducing RSV burden.

**Conclusions:** Our study is the first to comprehensively analyse hybrid RSV immunisation programs that use Abrysvo and nirsevimab. RSV immunisation programs can substantially reduce the burden of RSV in young children. Our modelling analysis provides evidence on immunisation type, timing, coverage, high-risk groups and dosage cost that will support decision makers and can be used in economic evaluations.

## Introduction

Respiratory syncytial virus (RSV) is a leading cause of respiratory infection worldwide, infecting people of all ages but with more severe outcomes in young children and the elderly [1–4]. The highest incidence of severe disease is in infants younger than 6 months. The incidence then decreases with age [5–7] with a resurgence of severity in adults over 65 years [3,4]. Recent global estimates report that the hospitalisation rate for RSV-associated acute lower respiratory infection (ALRI) peaks in children aged 0–3 months, and 39% of RSV-associated ALRI hospitalisations occur in the first 6 months of life [2]. In infants aged 0-6 months, those born preterm are at greatest risk for severe RSV illness and complications [8].

In recent years, new discoveries have resulted in two RSV-prevention products being licensed and implemented in programs globally as well as in Australia. Both the maternal vaccine, Abrysvo (Pfizer), and a long-acting humanised monoclonal antibody (mAb), nirsevimab (Beyfortus; Sanofi), provide passive immunisation to infants and young children. Abrysvo acts through the transplacental transfer of RSV-specific antibodies from the mother to the fetus during pregnancy while nirsevimab provides almost immediate protection directly to infants from ready-made monoclonal antibodies. Due to earlier licensing and government approvals granted, nirsevimab immunisation programs were in place before Abrysvo in several European countries and the USA over the northern hemisphere winter 2023/2024, and programs in some states in Australia over the southern hemisphere in winter 2024. The real-world effectiveness studies of nirsevimab in these countries indicate high effectiveness against hospitalisation ranging from 70-90% [9–13], with an estimated effectiveness of 88.2% [14] in preventing RSV-hospitalisation in infants and children under 2 years old in Western Australia (WA), where the most comprehensive nirsevimab program was implemented in Australia.

In November 2024, the Australian Federal Health Minister announced a hybrid RSV prevention program for 2025, with Abrysvo on the National Immunisation Program (fully funded by the federal government), and nirsevimab to be offered by individual States and Territories [15]. Australian States and Territories have rolled out very similar hybrid RSV immunisation programs for infants in 2025 [16–22]. Like the other Australian programs, the 2025 WA infant program focuses on providing the maternal vaccine to pregnant women at 28 to 36 weeks gestation (from 3rd February 2025) and offering nirsevimab to all infants at birth (from 1 April - 30 September 2025) if their mother did not receive the maternal vaccine. Nirsevimab is also offered, regardless of maternal vaccination status, to infants born to immunocompromised mothers, infants with pre-specified high-risk medical conditions, and those infants who were born less than 2 weeks from the mother receiving the vaccine [23]. In addition to the WA infant program, nirsevimab is offered to children entering their second RSV season with pre-specified medical conditions that place them at an increased risk for severe RSV infections. These include children born preterm, those with specific underlying medical conditions including chronic cardiac, respiratory, neurological, immunosuppressive or genetic/metabolic conditions, and all Aboriginal and Torres Strait Islander children. Although RSV prevention programs are now in place in Australia, there is a need for data-driven evidence to inform decisions on future programs with consideration to timing, coverage, effectiveness and cost of immunisation products [24]. The need for modelling evidence in different settings for the use of RSV immunisation products has been recognised as a priority in the WHO position paper published in May 2025 [25]. In this setting, models are needed to capture timing of interventions relative to the RSV season and the impact of hybrid programs.

In this study, we extend a dynamic compartmental model of RSV transmission in temperate WA [26,27] to incorporate both nirsevimab and Abrysvo immunisation, allowing a comprehensive analysis of potential RSV immunisation programs. We consider single-product and hybrid programs using both immunisations, timing of programs (either seasonal or year-round), differences in coverage levels and consider programs that target subgroups at higher risk of severe RSV, such as preterm infants. We also provide preliminary analysis of total dose cost associated with each program assuming dose costs of nirsevimab and Abrysvo based on the recent US Centre for Disease Control (CDC) Vaccine Price List [28]. There have been other model-based studies [29,30], some that explore possible impact before or early in the development of the most recent products [31–36], others that explore single-product RSV programs using nirsevimab [27,37,38], Abrsyvo [39] or directly compare single-product strategies of both [40,41]. Few of these studies model the Australian setting [27,32,39] and to date there are no modelling studies that comprehensively analyse hybrid immunisation strategies.

## Methods

### Setting and population-based data

WA is the largest state by area in Australia, and has a population of almost 3 million, mostly residing near the capital city of Perth in the southwest. RSV seasonality varies across the state with the changing climatic regions. In the temperate climate region, which includes metropolitan Perth and surrounds, RSV epidemics typically occur annually, peaking in the winter months (June to August). The RSV transmission model used in this study and described in [27], is calibrated to a time series of monthly RSV-hospitalisations in children aged <5 years in temperate WA (excluding the Pilbara and Kimberley regions) from 1 January 2015 to 31 December 2019 (see Figure S1) using the WA Respiratory Infections Linked Data Platform [42]. The WA Respiratory Infections Linked Data Platform is explained in [42] but in brief, it is a population-based cohort study, consisting of >350,000 WA births between 1 January 2010 and 31 December 2020, formed through individual level linkages of databases from the WA Department of Health including the Birth and Death Registry, Midwives Notification System, Hospital Morbidity Data Collection and PathWest Respiratory Virus Surveillance Dataset amongst other administrative datasets. An RSV-hospitalisation is defined as a hospital admission for any diagnosis where RSV was tested and found positive through polymerase chain reaction (PCR) with a respiratory specimen collection date within 4 days either side of the hospital admission date. RSV-hospitalisations were further separated into those children who were born preterm (defined as gestation week at birth less than 37 weeks) and those born term (defined as gestation week at birth 37 weeks or more). The resulting time series was of RSV-hospitalisations for children born preterm and term between 2015 and 2019 aggregated to a monthly time period (Figure S1).

### Base model structure and assumptions

The base model of RSV transmission has been described previously [26,27]. The model is a deterministic compartmental model of the Susceptible-Exposed-Infectious-Recovered-Susceptible form. The model is age-structured, dividing the population into 75 age groups (one-month age groups for children under 5 years and 5-year age groups thereafter) and captures the progression from first RSV exposure to repeat exposures, reducing infectivity after first exposure (Figure 1). The model is stratified to reflect a difference in risk of RSV-hospitalisation once infected in infants born full-term (≥37 weeks gestational age) compared to those born preterm (<37 weeks). An age-to-risk function formulated through fitting the model to hospitalisation time series data is used to translate RSV infections to hospitalisations by this risk grouping as well as age, with risk of hospitalisation scaled to decrease after the first infection (see Supplementary material).

**Figure 1:**
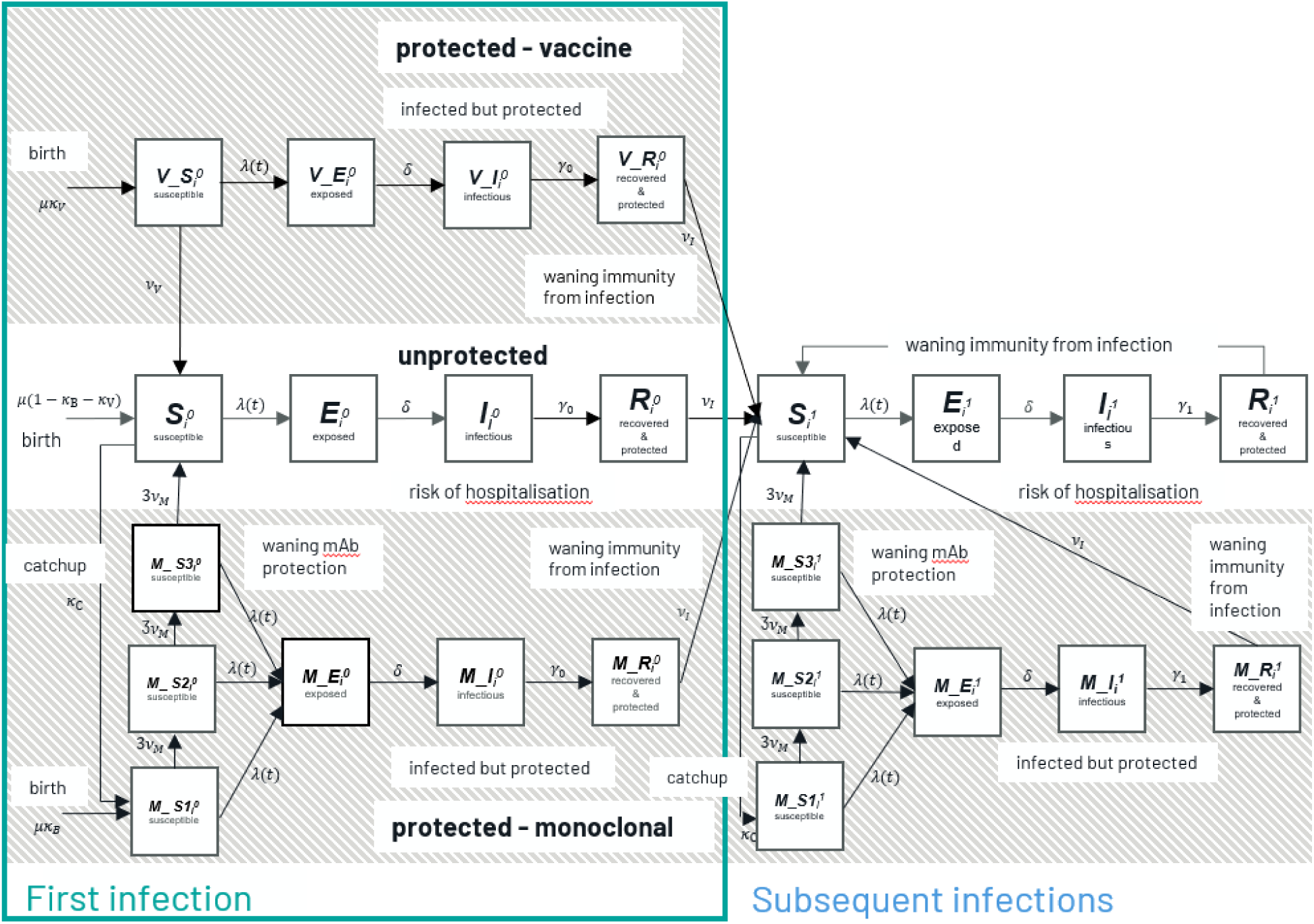
Schematic representation of the RSV transmission model including maternal vaccine, Abrysvo and monoclonal antibody, nirsevimab immunisation (at birth and older) for each age class i, where each state (susceptible, exposed, infectious, and recovered) for both naive (superscript 0) and repeat (superscript 1) exposures, represents a proportion of the total population. 𝝀(t) represents the force of infection, which is driven by the proportion of the population that is infectious and incorporates seasonally-fluctuating transmission, 1/δ is the latency period, 1/ɣ_0_ and 1/ɣ_1_ are the infectious periods for first and subsequent infections, 1/𝜈_I_ is the immunity period after infection and 1/𝜈_M_ the immunity period after mAb immunisation. The birth rate is denoted by μ, and Κ_B_ and Κ_C_ are the mAb coverage rates for newborns and older infants respectively, and Κ_V_ is the maternal vaccine coverage rate.

The model assumes that newborns of mothers who have been recently infected with RSV have some protection resulting from a transplacental transfer of RSV antibodies during the third trimester that wanes over the first year of life [43]. We simulate the seasonal effect in transmission, corresponding to an increase in activity in the winter months in temperate WA, by use of a sinusoidal forcing term with the associated amplitude and transmission coefficients calibrated to the hospitalisation data in model fitting. Though RSV seasonality changed in WA and elsewhere during the COVID-19 pandemic [44], viral activity has returned to the pre-COVID-19 norm [45] that is captured in this model.

The underlying demographics are based on the 2016 Australian Bureau of Statistics (ABS) population for WA, minus the subtropical and tropical northern regions (Kimberley and Pilbara) as defined by postcode [46], consistent with the 2015-2019 RSV-hospitalisation time series used for model calibration. A synthetic contact matrix specific to temperate WA, capturing the probability of infection due to contact mixing between age groups, was constructed using the R package *conmat* [47–49], the 2016 ABS 5% microdata sample for the Greater Perth region [50], and the Childhood Education and Care Survey 2017 [51]. Further details on model assumptions, fitting and parameterisation can be found in the Supplementary material.

### Immunisation model structure and assumptions

In [27] we describe the extension of the model to simulate the administration of nirsevimab to different age and risk groups. We assume that nirsevimab does not affect the risk of RSV acquisition or transmission [52], and that only the severity of RSV disease is reduced.

Nirsevimab protection was simulated in the model by an Erlang-3 distribution with an average of 78% protected from RSV-hospitalisation over the 150 days following immunisation (Figure S8). This is consistent with clinical trial data [53] and real-world effectiveness studies [9–14].

In this study, we further extend the model through the addition of compartments to simulate the protection of newborns whose mothers received the maternal vaccine, Abrysvo (Figure 1). An exponential waning profile consistent with the efficacy estimates of Abrysvo against RSV-hospitalisation in clinical trials [54] is used to simulate the protection characteristics of Abrysvo in the model (Figure S9). Figure S10 compares the protection waning profiles of nirsevimab and Abrysvo, showing more sustained protection and hence higher efficacy of nirsevimab over the first 150 days. We assume here that, as with nirsevimab, Abrysvo affects only the severity of infection and has no impact on acquisition or transmission when protecting the infant as current evidence to suggest otherwise is limited, and the potential herd effect of vaccinating mothers has been reported as minimal in other modelling studies [31–33,39,55].

As indicated in Figure 1, the model allows for the immunisation of multiple cohorts of infants and young children. Though Abrysvo only protects mothers and their newborns, nirsevimab can be administered to older infants as well as at birth. The model accommodates scenarios that apply to multiple cohorts of children. For example, the 2024 WA RSV program includes infants immunised at birth (‘newborn cohort’), older infants entering their first RSV season (‘catch-up cohort’), and eligible children with pre-specified high-risk conditions (‘high-risk group’, which for this study was the preterm risk group defined as in the model assumptions) entering their second RSV season. Additional assumptions concerning older infants and young children are that those in the catch-up and high-risk cohorts receiving nirsevimab prior to their second RSV season were immunised in advance of the season (April - May). Immunisation only occurs in the model to eligible children who are not currently infected, do not have temporary immunity from a recent RSV infection, or are not already protected by RSV immunisation. Although predominately protection to infants was through either maternal vaccination or post-natal nirsevimab, in select situations in the 2025 WA infant program, a mother could be immunised with Abrysvo and the newborn could also be eligible for nirsevimab, hence having both immunisations for the same birth. We assume that, as eligibility in this case directly relates to concern that the maternal vaccine is not as effective, the overriding protection to the infant comes from nirsevimab. As we do not yet have data as to the number of newborns who have received nirsevimab at birth after their mother has received Abrysvo, we have not adjusted dose counts to reflect this. Therefore, in this study, coverage of Abrysvo relates to the proportion of infants who were only protected by Abrysvo at birth.

### Immunisation scenarios

The RSV immunisation programs modelled are shown in Table 1. We consider Abrysvo-only and nirsevimab-only programs where a dose is administered at birth, as well as a nirsevimab program replicating the 2024 WA program that applies to multiple cohorts. These scenarios capture different assumptions of timing, either year-round or seasonal, where protection is provided only in April-September when RSV activity is at its highest in temperate WA. Coverage is assumed to be either 50%, 70% or 90%, except for the 2024 WA program scenario that replicates the published 2024 WA coverage levels [56].

**Table 1:** The immunisation program scenarios modelled and analysed. Nirsevimab-only (mAb), Abrysvo-only (mat vac) and hybrid programs. Seasonal immunisation occurs from April to September.

|  | Scenario |  | Coverage |
| --- | --- | --- | --- |
| **mAb** | 1 | **Year-round at birth** | (a) 50% (b) 70% (c) 90% |
|  | 2 | **Seasonal at birth** | (a) 50% (b) 70% (c) 90% |
|  | 3 | **Seasonal at birth with catch-up + 2nd season high-risk (2024 WA program)** | birth: 79%<br>catch-up: 65%<br>2nd season high-risk: 30% |
| **mat vac** | 4 | **Year-round** | (a) 50% (b) 70% (c) 90% |
|  | 5 | **Seasonal** | (a) 50% (b) 70% (c) 90% |
| **hybrid** | 6 | **Year-round mat vac + seasonal mAb at birth high-risk** | mat vac: (a) 50% (b) 70% (c) 90%<br>mAb: 100% (high-risk) |
|  | 7 | **Year-round mat vac OR seasonal mAb at birth** | mat vac: (a) 30% (b) 45% (c) 60%<br>mAb: (a) 50% (b) 35% (c) 20% |
|  | 8 | **Year-round mat vac OR seasonal mAb at birth + 2nd season mAb high-risk** | mat vac: (a) 30% (b) 45% (c) 60%<br>mAb (at birth): (a) 50% (b) 35% (c) 20%<br>mAb (2nd season high-risk): 30% |
|  | 9 | **Year-round mat vac OR seasonal mAb at birth + catch-up + 2nd season mAb high-risk (2025 WA program)** | mat vac: (a) 30% (b) 45% (c) 60%<br>mAb (at birth): (a) 50% (b) 35% (c) 20%<br>mAb (catch-up): 70%<br>mAb (2nd season high-risk): 30% |

The four hybrid programs combine the use of Abrysvo and nirsevimab with differing focus. Scenario 6 provides year-round access to Abrysvo at varying coverage levels with nirsevimab reserved for the protection of high-risk infants at birth. This scenario reflects the possibility that nirsevimab may be significantly more expensive than Abrysvo [28] and hence reserved for those at highest risk. A similar approach, though reserved for a more restricted target population, was implemented in previous programs using palivizumab. Palivizumab is a short-acting monoclonal antibody approved for use in Australia in 1999 [57] and due to its high cost, was only offered to a very select group of infants considered at high-risk of RSV due to comorbidities or premature birth [58,59].

Scenarios 7, 8 and 9 reflect hybrid programs where the maternal vaccine is offered year-round but during the temperate RSV season, there is the choice of Abrysvo vaccination of the mother or immunisation of infants at birth with nirsevimab. Scenarios 8 and 9 also target high-risk children entering their second RSV season and scenario 9 additionally provides an opportunity for a “catch-up” dose for those infants entering their first RSV season that were either born outside of the RSV season or were not protected by either immunisation type at birth. Scenario 9 most closely matches the WA program being implemented in 2025. The coverage levels for Abrysvo and nirsevimab considered in scenarios 7, 8 and 9 were chosen to reflect an overall newborn coverage of 80% during the RSV season, consistent with the coverage obtained for the birth cohort in the 2024 WA RSV program [56]. As the acceptance of maternal vaccination varies according to the antigen, program, and over time [60], a range of scenarios were considered ranging from 30% to 60% coverage of Abrysvo, with seasonal nirsevimab coverage at birth ranging from 20-50%.

For each scenario modelled, we calculate the number and proportion of annual RSV-hospitalisations averted in temperate WA when compared to model predicted RSV-hospitalisations assuming no immunisation. These results are presented by age; under 3 months, between 3 and 6 months, between 6 and 12 months, and under 24 months. We also calculate the number of doses of each immunisation type administered under each scenario and use this and the number of RSV-hospitalisations averted to calculate the number needed to immunise (NNI) to avert one RSV-hospitalisation.

### Cost of dose

In Australia, the price of immunisation products purchased by the government for national and state programs is not publicly available. Unlike nirsevimab, Abrysvo is also available on the Australian private market at $331.99 AUD per dose [61]. The US CDC Vaccine Price List of May 2025 [28] indicates a CDC cost per dose of $414.75 US for 50mg or 100mg nirsevimab (Beyfortus) and $230.09 US for Abrysvo. Based on US prices, we looked at three scenarios of dosage cost of nirsevimab relative to Abrysvo. We considered nirsevimab and Abrysvo equal in cost at $200 US per dose, nirsevimab cost 1.5 times that of Abrysvo at $300 US per dose, and nirsevimab double the cost of Abrysvo at $400 US per dose. Using these price points, we calculated the annual dosage cost of each of the modelled immunisation program scenarios per 100,000 children under the age of 2-years-old. This simple dose cost analysis is not a cost-effectiveness analysis but a preliminary exploration of the implications of a potential significant dose cost differential between the two preventative products.

## Results

### Single-product programs

For the same assumed coverage and target population, the nirsevimab-only programs avert more RSV-hospitalisations and are more efficient in reducing RSV-hospitalisation burden (Figure 2, Table 2). A year-round nirsevimab program at 70% coverage is estimated to avert 33% of RSV-hospitalisations of under 2-year-olds annually, including averting 60% of RSV-hospitalisations of under 3-month-olds, the most vulnerable age group to severe RSV infections. In comparison, a year-round Abrysvo program with the same coverage is estimated to avert 25% of annual RSV-hospitalisations of under 2-year-olds including 48% of RSV-hospitalisations associated with under 3-month-olds. For both single-product scenarios, seasonal programs that focus on protecting infants in the 6 months of the year where RSV activity is the highest (ie. seasonal programs) are most efficient. The NNI was 88 for a seasonal program compared to 113 for a year-round nirsevimab program; and 111 for a seasonal program compared to 147 for a year-round Abrysvo program (Table 2). Though seasonal programs are a more efficient use of immunisation doses, they are less effective for the older age groups. Year-round programs have a proportionally higher impact on the older age groups than the youngest age group. For example, nirsevimab at 70% coverage for the full year only increases impact on under 3-month-olds from 46% to 60%, whereas the impact on 3 to 6-month-olds increases by more than double from 17% to 35%, and by more than triple for the 6 to 12-month-olds from 4% to 13% (Table 2).

**Figure 2:**
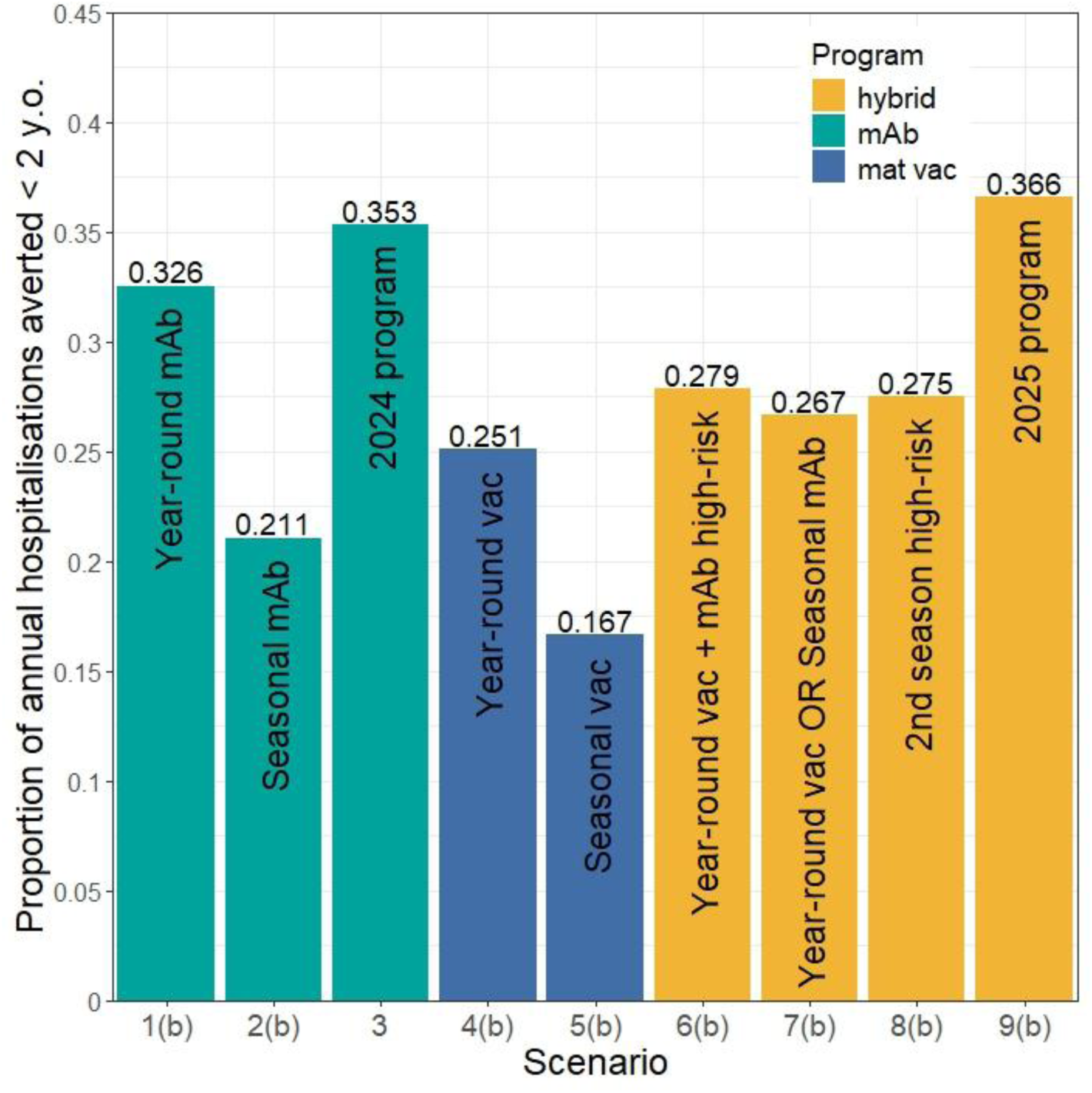
Proportion of annual RSV-hospitalisations averted of under 2-year-olds in temperate WA for nirsevimab-only (mAb)(Scenarios 1-3), Abrysvo-only (mat vac) (Scenarios 4-5) and hybrid immunisation programs (Scenarios 6-8). For the year-round and seasonal nirsevimab and Abrysvo programs, the 70% coverage scenario results are shown. For the hybrid programs, “Year-round vac + mAb high-risk” corresponds to year-round 70% Abrysvo coverage with mAb for all high-risk infants, “Year-round vac OR Seasonal mAb”, “2nd season high-risk” and “2025 program” correspond to year-round 45% Abrysvo coverage and seasonal 35% nirsevimab coverage.

**Table 2:** Summary of a subset of model scenario results. Values presented are the proportion of hospitalisations averted yearly for under 3-month, 3-to-6 month, 6-to-12 month, and under 24-month children, the number of yearly nirsevimab (mAb) and Abrysvo (mat vac) doses associated with a program and the number needed to immunise (NNI) to prevent one RSV-hospitalisation. See Table S5 for full results.

|  | Scenario and coverage |  | Prop. of hosp. averted |  |  |  | Number of doses |  |  | NNI |
| --- | --- | --- | --- | --- | --- | --- | --- | --- | --- | --- |
|  |  |  | <3m | 3-<6m | 6-<12m | <24m | mAb | mat vac | total |  |
| **mAb** | 1(b) | **Year-round at birth 70%** | 0.60 | 0.35 | 0.13 | 0.33 | 21533 | 0 | 21533 | 113 |
|  | 2(b) | **Seasonal at birth 70%** | 0.46 | 0.17 | 0.04 | 0.21 | 10766 | 0 | 10766 | 88 |
|  | 3 | **Seasonal at birth with catchup + 2nd season high-risk (2024 WA program)**<br>79% birth, 65% catch-up, 30% high-risk | 0.56 | 0.39 | 0.24 | 0.35 | 19389 | 0 | 19389 | 94 |
| **mat vac** | 4(b) | **Year-round 70%** | 0.48 | 0.24 | 0.10 | 0.25 | 0 | 21533 | 21533 | 147 |
|  | 5(b) | **Seasonal 70%** | 0.37 | 0.12 | 0.03 | 0.17 | 0 | 10766 | 10766 | 111 |
| **hybrid** | 6(b) | **Year-round mat vac + seasonal mAb at birth high-risk**<br>70% mat vac, 100% mAb high-risk | 0.53 | 0.27 | 0.11 | 0.28 | 1415 | 20542 | 21957 | 135 |
|  | 7(b) | **Year-round mat vac OR seasonal mAb at birth**<br>45% mat vac, 35% mAb | 0.54 | 0.24 | 0.09 | 0.27 | 5383 | 13843 | 19226 | 123 |
|  | 8(b) | **Year-round mat vac OR Seasonal mAb + extra high-risk**<br>45% mat vac, 35% mAb, 30% high-risk | 0.54 | 0.24 | 0.10 | 0.28 | 5919 | 13843 | 19761 | 123 |
|  | 9(b) | **Year-round mat vac OR seasonal mAb at birth + catchup + 2nd season mAb high-risk (2025 WA program)**<br>45% mat vac, 35% mAb at birth, 70% catch-up, 30% high-risk | 0.57 | 0.41 | 0.27 | 0.37 | 11690 | 13843 | 25532 | 120 |

**Figure 3:**
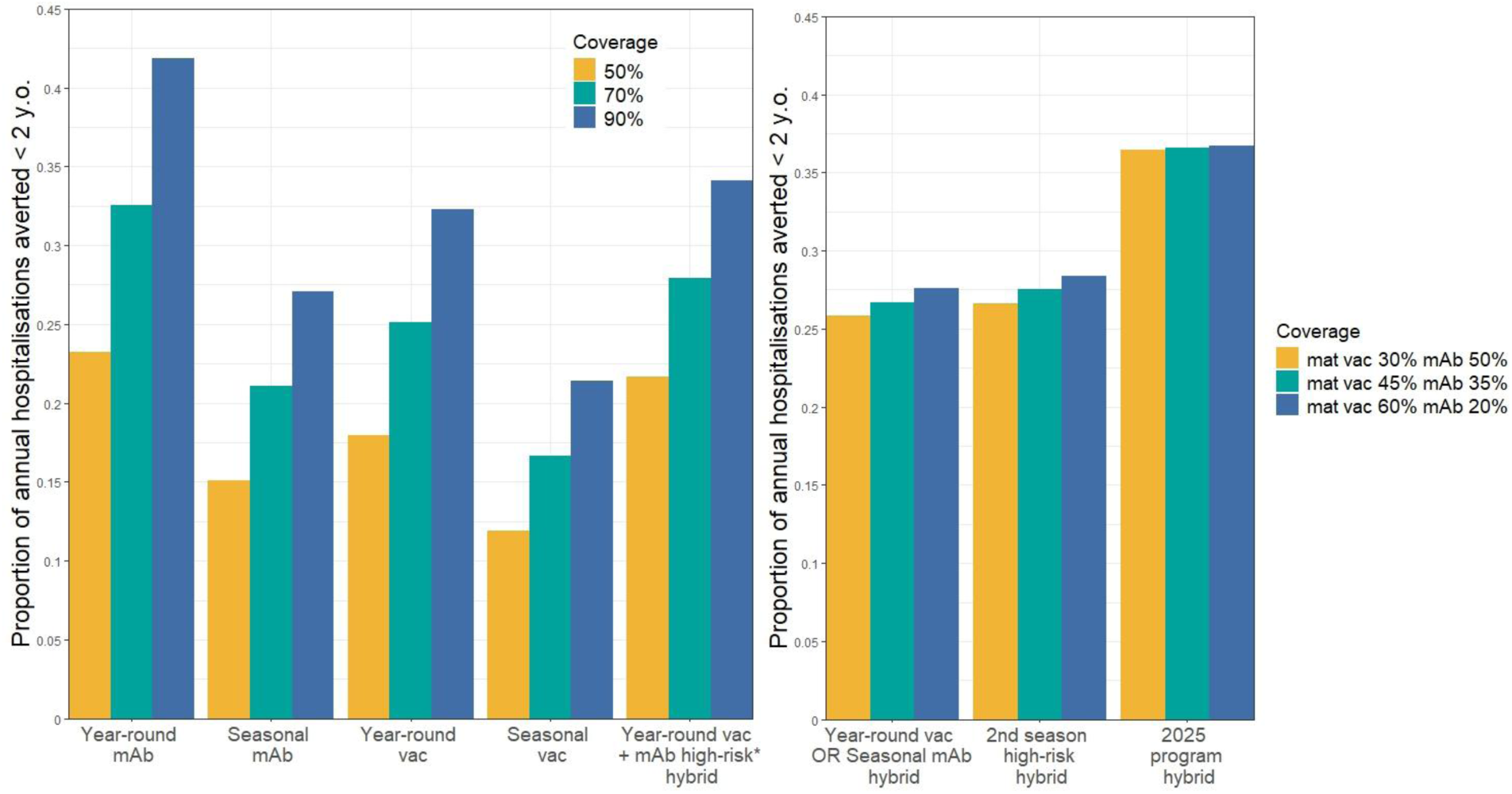
Proportion of annual RSV-hospitalisations averted in under 2-year-olds in temperate WA under different assumptions of coverage for potential immunisation programs. Scenarios shown here correspond to mAb scenarios 1 and 2, mat vac scenarios 4 and 5, and hybrid scenarios 6, 7, 8 and 9. *For the hybrid scenarios, the colour of the bar relates to the maternal vaccine coverage. See Table 1 for full descriptions of scenarios.

### Hybrid programs

The hybrid program offering year-round maternal vaccination to all pregnant mothers and nirsevimab immunisation at birth for infants at increased risk of severe RSV (scenario 6(b)), has higher impact and efficiency than the year-round Abrysvo-only scenario (scenario 4(b)) due to the targeted approach for high-risk infants (NNI of 135 compared to 147, see Table 2).

Increased use of nirsevimab in the three other hybrid programs (scenarios 7(b), 8(b) and 9(b)) translates to more effective, and in some cases more efficient, immunisation programs than those relying only on Abrysvo. Despite requiring over 1500 less doses overall, scenario 8(b) offering the maternal vaccine year-round (at 45% coverage) with nirsevimab offered seasonally (at 35% coverage) and an additional second season dose for high-risk infants (30% coverage), has a higher impact on RSV-hospitalisations averted (54% for under 3 months, 28% for under 2 years) than a year-round Abrysvo-only program at 70% coverage (scenario 4(b), 48% for under 3 months, 25% for under 2 years).

### Coverage

For each of the single-product programs, and the hybrid program with year-round Abrysvo and nirsevimab only offered to high-risk infants, an increase in coverage from 50% to 90% provides a corresponding significant reduction in RSV burden. For example, the year-round nirsevimab scenario at 90% coverage averts 42% of annual RSV-hospitalisations in under 2-year-olds compared to 23% at 50% coverage, and the seasonal Abrysvo scenario at 90% coverage averts 21% of annual RSV-hospitalisations in under 2-year-olds compared to 12% at 50% coverage.

Hybrid scenarios 7, 8 and 9 illustrate the impact of increasing year-round coverage of the maternal vaccine while decreasing newborn seasonal coverage of nirsevimab, while maintaining a seasonal combined coverage of 80% for newborns. There is a small increase in impact on annual averted RSV-hospitalisations as year-round coverage levels of Abrysvo increase in scenarios 7 and 8, but this increase in the year-round maternal vaccine coverage has minimal impact for scenario 9. The small increase for scenarios 7 and 8 corresponds to the proportionally higher impact of year-round coverage on the older age groups, and this is not as evident in scenario 9 as the inclusion of the catch-up program has the overriding impact on older infants that were born outside of the high RSV activity months. The hybrid scenarios 7-9 that have higher seasonal nirsevimab coverage have higher efficiency, evidenced by lower NNI (see Table S5), for a relatively small difference in reduction of burden to hybrid programs with high year-round Abrysvo coverage.

### Dose cost scenarios

For equal cost of nirsevimab and Abrysvo at $200 US per dose, the 2025 WA program scenario has the highest annual dosage cost per 100,000 children under 2-years-old at close to $8.5 million US (Figure 4, Table 2) and the highest reduction in RSV-hospitalisation burden, impacting not only the youngest age group but also infants at the older age range (Figure 2, Table 2). At $400 US for nirsevimab and $200 US for Abrysvo, the annual dosage cost of the 2025 program rises to close to $12 million US per 100,000 children under 2-years-old (Figure 4). However, this is less than the year-round nirsevimab and 2024 WA nirsevimab program, despite requiring a larger number of doses (Table 2). Figure 4 illustrates that if nirsevimab costs more than Abryso, the annual dosage cost of hybrid programs increases at a slower rate than nirsevimab-only programs while being highly effective at reducing RSV burden (Figure 2).

**Figure 4:**
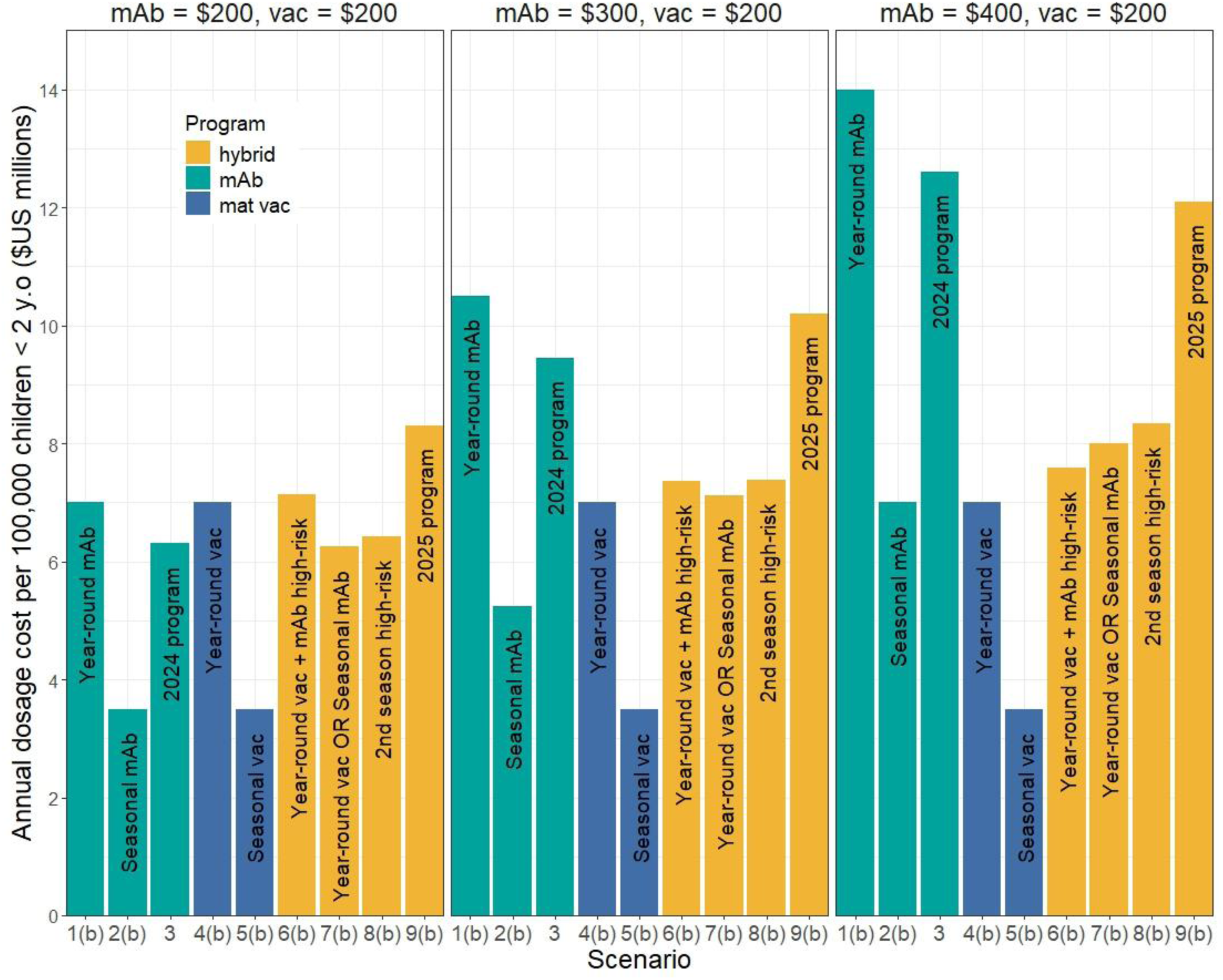
Annual dosage cost per 100,000 children under 2 years old for each of the modelled scenarios under the immunisation unit costs assumed. The left panel assumes equal dose cost of nirsevimab and Abrysvo at $200 US, the middle panel assumes nirsevimab is 1.5 x the dose cost of Abrysvo or $300 US, and the right panel assumes nirsevimab is double the dose cost of Abrysvo or $400 US.

## Discussion

In this study, we estimate the impact of a set of RSV immunisation programs with consideration to immunisation type, timing of administration, coverage levels and target cohorts, in the context of temperate WA RSV epidemiology. We find that all programs analysed were effective in reducing the burden of severe RSV in infants and young children. Due to the product characteristics of nirsevimab, programs using nirsevimab for newborn infants at similar coverage levels to the Abrysvo programs, averted more RSV-hospitalisations annually. Seasonal programs that focused on protection during high RSV activity and programs targeting high-risk infants were the most efficient relating to the lowest values of NNI. When dose cost scenario results are analysed alongside scenario impact on RSV burden, we find that hybrid programs are not only highly effective in averting RSV-hospitalisations in under 2-year-olds but, depending on the dosage cost of the two immunisation products, could balance the potential higher dose cost of nirsevimab with nirsevimab’s higher product effectiveness. Hybrid programs, like nirsevimab-only programs, also have the advantage of targeting multiple age cohorts and high-risk groups. This helps to provide equitable access to RSV immunisation, particularly across birth months where, for example, infants born outside of the RSV season in temperate climatic regions could potentially enter their first RSV season with no immunisation protection due to waning, despite being protected at birth.

The availability of two effective immunisation products for the protection of infants and young children from severe RSV has the potential to greatly reduce the future burden of RSV globally, but there is still a lot of uncertainty as to how to use these products optimally. Studies like ours can fill this evidence gap, enabling full consideration of hybrid programs and timing relative to the RSV season [24,25]. RSV infant immunisation programs in place over the 2024/25 northern hemisphere winter have varied with Italy and Spain, for example, continuing with a nirsevimab program, the UK favouring a maternal vaccine program, and the USA and France, like Australia, implementing a hybrid strategy [62–66]. Other countries, like New Zealand, have yet to implement an RSV infant immunisation program that takes advantage of the new immunisation products currently available [67]. A third product, clesrovimab, also a long-acting monoclonal antibody but with the benefit of only requiring a single dose regardless of infant weight, has achieved high efficacy against RSV-hospitalisation in clinical trials and was recently approved for use in the US [68,69]. The availability of multiple products has the advantage of helping with supply issues and encouraging competitive pricing [70], but can also require more complex decision-making. This decision process can happen at the government or implementation level, or may, in a hybrid program, be partially pushed onto the individual. In this case, parental preference of product-type has importance in the development of an immunisation program, and parents may have more reason to look for reliable evidence to guide decisions than if there were only one immunisation option [71]. This study not only can be used to inform policy development but can underpin individual decision-making as to the benefits of one immunisation type over another and timing of administration depending on month of birth, for example.

Our study is not without limitations. As previously described [27], this model has only been calibrated to temperate WA RSV dynamics and so analysis of immunisation strategies in northern WA, where RSV activity does not have a clearly defined peak season, is the subject of future work. This study also considers only preterm infants in the “high-risk” group, though higher risk of severe RSV is also associated with Aboriginal children and infants with comorbidities such as chronic respiratory diseases, congenital heart disease and immunodeficiency or immunosuppressive conditions [42]. Data for high-risk groups are particularly difficult to attain due to lack of accurate comorbidity data in the immunisation register to allow identification of those children. This makes it harder to ascertain accurate coverage levels for these subgroups. The scenarios analysed in this study emphasise the benefit of prioritising immunisation to preterm infants who disproportionately suffer the burden of RSV.

This study includes three dose cost scenarios for nirsevimab price relative to Abrysvo to illustrate a key factor likely to influence any comparison of single-product and hybrid RSV immunisation programs by policy makers. There is a clear need for further economic analysis. A cost-effectiveness analysis, such as the one done by Hodgson *et al* [41], but extended to analyse hybrid programs and in the Australian setting could build on the results in this paper to provide crucial evidence for policy makers. As real-world effectiveness estimates are increasingly made available, the model can be updated and used to re-evaluate immunisation scenarios. Similarly, as data becomes available as to the number of infants receiving nirsevimab at birth after their mother has received Abrysvo, as well as the relative cost of nirsevimab versus Abrysvo, the model can be used to optimise the balance of Abrysvo to nirsevimab usage in a hybrid program.

Our findings are consistent with other modelling studies when comparing similar scenarios [37,41], despite differences in model structure, assumptions and setting. Though three other modelling studies have included examples of hybrid programs [72–74], no modelling study, at time of publication, has comprehensively analysed hybrid programs combining Abrysvo and nirsevimab use. The need for this modelling work to help guide future decisions on the optimal combination of immunisation products has been recognised [24]. Our study provides much needed evidence to help meet this gap and to guide RSV immunisation policy development.

## Data availability

The datasets generated and/or analysed during the current study are not publicly available due to the terms of the ethics approval granted by the Western Australian Department of Health Human Research Ethics Committee and data disclosure policies of the Data Providers.

## Code availability

The R program code for the dynamic transmission model is available on *github* at https://github.com/fionagi/rsvmod.imm25

## Contribution

FG, ABH, KG and HCM conceptualised and designed the study. FG developed the model code and conducted analysis with technical input from ABH and KG. HCM advised on RSV epidemiology. CCB provided effectiveness and coverage estimates and input into the modelled scenarios. FG wrote the first draft. All authors contributed with critical input, reviewing and editing of the manuscript, and have approved the final version.

## Data Availability

https://github.com/fionagi/rsvmod.imm25

## Acknowledgements

The authors would like to thank the Linkage, Data Outputs and ISPD Client Services Teams at Western Australian Data Linkage Services, as well as the Western Australia Midwife Notification System, Emergency Department Data Collection, Hospital Morbidity Data Collection, Birth and Death Registrations Data Collections, WA Notifiable Infectious Diseases Database, and PathWest Laboratory Medicine Database.

The STAMP Investigator Team is Hannah C Moore, Christopher C Blyth, Samantha Carlson, Fiona Giannini, Mohinder Sarna, David Foley, Catherine Hughes, Peter Richmond, Avram Levy, Ewan Cameron

Stan Perron Charitable Foundation (STAMP Program, ref: 00046ProgPart)

