## Supplementary material for "Modelling the impact of long-acting monoclonal antibody, maternal vaccine and hybrid programs of RSV immunisation in temperate Western Australia"

Hospitalisation data for under 5-year-old children	2

Parameter values	3

Model equations	5

Contact matrix	9

Model fitting	10

Age-to-risk function	10

Log likelihood	10

Parameter estimates	11

Model fit of hospitalisation predictions to observed data	14

Age-to-risk	17

Modelled proportion of mothers passing on antibodies	18

Immunisation protection profiles	19

mAb	19

Maternal vaccine	20

Scenario output	22

**References	2**7

### Hospitalisation data for under 5-year-old children

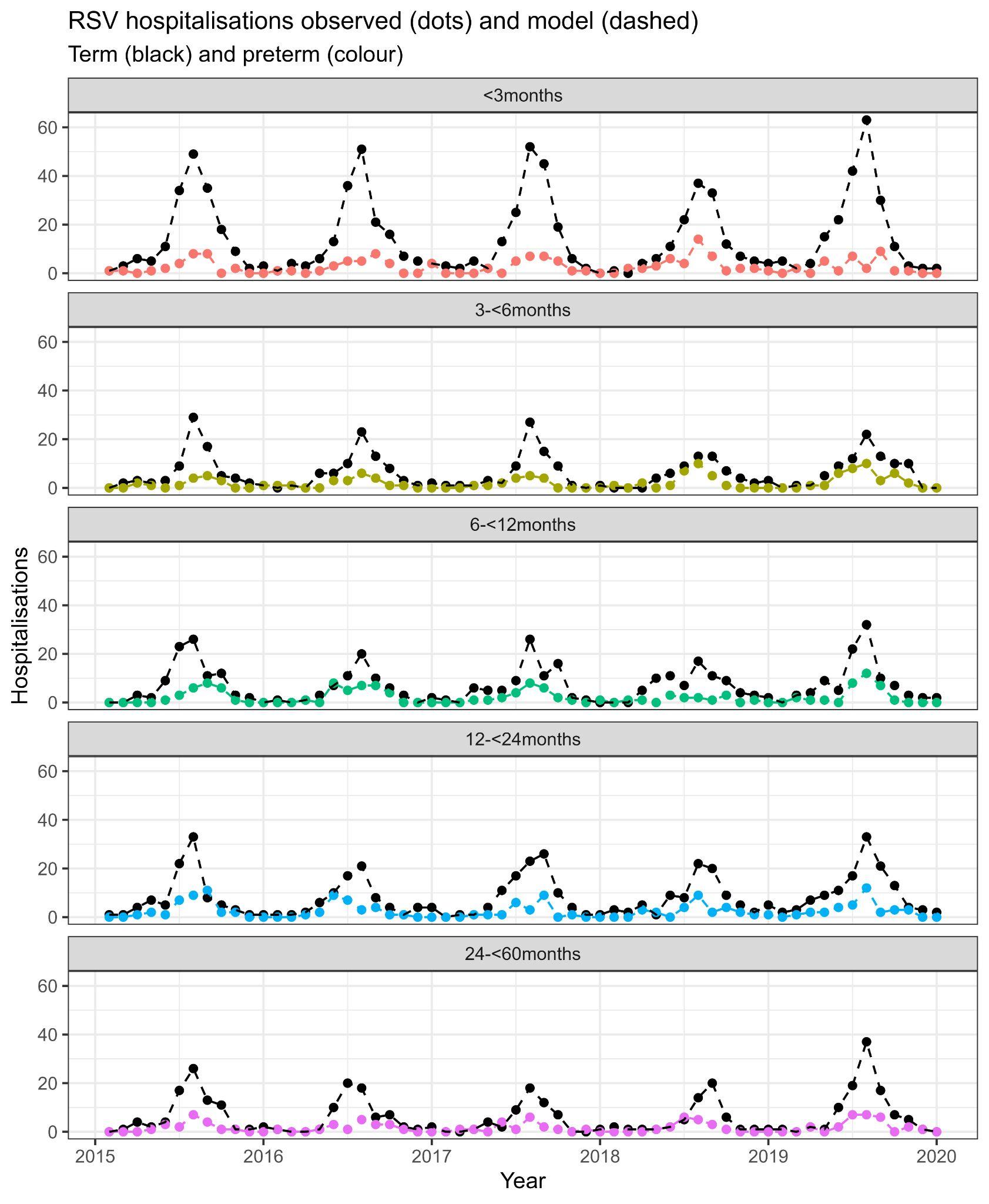

**Figure S1:** Monthly RSV-hospitalisations in temperate WA for children under 5 years old, divided into infants born term (black dots) and infants born preterm (coloured dots).

### Parameter values

**Table S1**: Parameter values for base and risk models.

| **Parameter** | **Definition** | **Fixed/Fitted** | **Value(s)** | **Reference** |
| --- | --- | --- | --- | --- |
| 1/ɣ_0_ | Infectious period (days) for first exposure | Fixed | 10 | [[1,2]](https://paperpile.com/c/Y68o4Y/lU8aQ+PR0cj) |
| 1/ɣ_1_ | Infectious period (days) for second (and subsequent) exposures | Fixed | 7 | [[1,2]](https://paperpile.com/c/Y68o4Y/lU8aQ+PR0cj) |
| ῶ | Reduced infectiousness of those who have experienced at least one prior infection | Fixed | 0.7 | [[1,3,4]](https://paperpile.com/c/Y68o4Y/lU8aQ+5EZx3+96rht) |
| 1/δ | Latent period (days) | Fixed | 4 | [[5]](https://paperpile.com/c/Y68o4Y/lsUt1) |
| 1/𝜈_I_ | Immunity period due to infection (days) | Fixed | 230 | [[6]](https://paperpile.com/c/Y68o4Y/CgArA) |
| 1/η | Durability of natural maternal immunity (days) | Fixed | 142.5 | [[7]](https://paperpile.com/c/Y68o4Y/b1QRn) |
| 𝜎_i_ | Reduced susceptibility due to natural maternal immunity at age, i where i is in months for infants less than 1 year old | Fixed | See Table S2 | [[7]](https://paperpile.com/c/Y68o4Y/b1QRn) |
| 𝜎_E_ | Reduced susceptibility due to previous exposure | Fixed | 0.77 | [[5,8–11]](https://paperpile.com/c/Y68o4Y/bCo1q+lsUt1+TlS5i+wcmCe+xwp0Q) |
| b_0_ | Transmission coefficient | Fitted | 0.0494 | Fitted value |
| b_1_ | Amplitude of seasonal forcing | Fitted | 0.1437 | Fitted value |
| A | Average maximum risk of hospitalisation of term infants (at age 0) | Fitted | 0.0260 | Fitted value |
| B | Decay constant | Fitted | 0.1852 | Fitted value |
| C | Average minimum risk of hospitalisation across all ages | Fitted | 0.0071 | Fitted value |
| D | Scaling factor lowering risk for those with prior exposures | Fixed | 0.2 | [[12,13]](https://paperpile.com/c/Y68o4Y/gSO2g+ZVBt8) |
| N | Total population of southern WA (ages 0-79) | Fixed | 2,288,349 | ABS population 2016 [[14]](https://paperpile.com/c/Y68o4Y/8cxJl) |
| 𝛂 | Proportion of births that are preterm (<37 weeks) | Fixed | 0.0917 | WA RSV linked data [[15]](https://paperpile.com/c/Y68o4Y/MPtN9) |
| E | Scaling factor increasing risk for preterm infants | Fitted | 2.5482 | Fitted value |
| 1/𝜈_M_ | mAb average durability (days) | Fixed | 150 | [[16–18]](https://paperpile.com/c/Y68o4Y/OWODD+8goIp+H2PRX) |
| 1/𝜈_v_ | Maternal vaccine average durability (days) | Fitted | 120 | Figure S9 |

**Table S2**: Reduced susceptibility due to natural maternal immunity (𝜎_i_) for the first year of life, assuming 37% of mothers pass on RSV antibodies at birth and protection decays exponentially with a mean of 1/λ = 142.5 days as in [[7]](https://paperpile.com/c/Y68o4Y/b1QRn). These are monthly midpoints.

| **Age** (months) | **𝜎_i_** |
| --- | --- |
| 0-<1 | 0.6674784 |
| 1-<2 | 0.7314309 |
| 2-<3 | 0.7830837 |
| 3-<4 | 0.8248023 |
| 4-<5 | 0.8584973 |
| 5-<6 | 0.8857119 |
| 6-<7 | 0.9076925 |
| 7-<8 | 0.9254456 |
| 8-<9 | 0.9397843 |
| 9-<10 | 0.9513653 |
| 10-<11 | 0.9607190 |
| 11-<12 | 0.9682738 |

### Model equations

The equations below describe the base and immunisation models respectively, where the base model does not include immunisation, and the immunisation model includes the use of mAb at different ages and the protection of the maternal vaccine at birth.
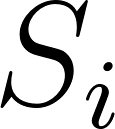
<https://www.codecogs.com/eqnedit.php?latex=S_i#0> represents the number of susceptible individuals in age group
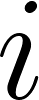
<https://www.codecogs.com/eqnedit.php?latex=i#0>,
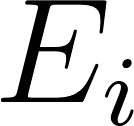
<https://www.codecogs.com/eqnedit.php?latex=E_i#0> represents the number of exposed individuals (infected but not yet infectious),
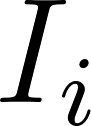
<https://www.codecogs.com/eqnedit.php?latex=I_i#0> represents the number of infectious individuals, and
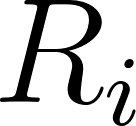
<https://www.codecogs.com/eqnedit.php?latex=R_i#0> represents the number of recovered and temporarily immune individuals. The superscripts 0 and 1 indicate naive and subsequent exposures respectively. The indices
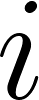
<https://www.codecogs.com/eqnedit.php?latex=i#0> and
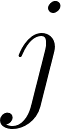
<https://www.codecogs.com/eqnedit.php?latex=j#0> represent the 75 age cohorts where
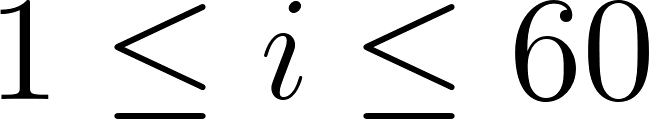
<https://www.codecogs.com/eqnedit.php?latex=1%20%5Cle%20i%20%5Cle%2060#0> are the monthly age groups from 0 to 59 months of age, and then
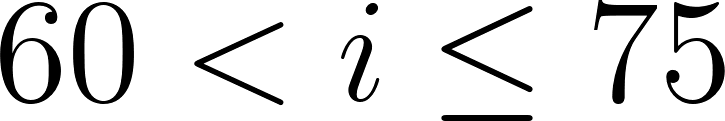
<https://www.codecogs.com/eqnedit.php?latex=60%20%3C%20i%20%5Cle75#0> are 5-year age groups starting at 5-9 years.

The transmission function
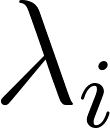
<https://www.codecogs.com/eqnedit.php?latex=%5Clambda_i#0> is the force of infection on age group
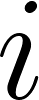
<https://www.codecogs.com/eqnedit.php?latex=i#0> at time
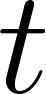
<https://www.codecogs.com/eqnedit.php?latex=t#0>. The transmission function phase shift parameter, ø, was fixed by the assumption that peak infections occurred in July, and the contact matrix
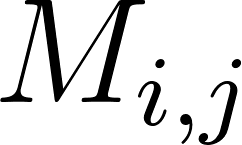
<https://www.codecogs.com/eqnedit.php?latex=M_%7Bi%2Cj%7D#0>, represents the number of contacts that an individual in age group
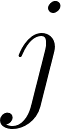
<https://www.codecogs.com/eqnedit.php?latex=j#0> has with individuals in age group
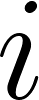
<https://www.codecogs.com/eqnedit.php?latex=i#0>. When
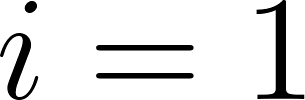
<https://www.codecogs.com/eqnedit.php?latex=i%3D1#0>,
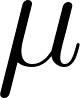
<https://www.codecogs.com/eqnedit.php?latex=%5Cmu#0> is the birth rate and
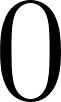
<https://www.codecogs.com/eqnedit.php?latex=0#0> otherwise. The ageing rate of individuals in age group
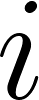
<https://www.codecogs.com/eqnedit.php?latex=i#0> is represented by
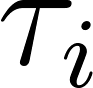
<https://www.codecogs.com/eqnedit.php?latex=%5Ctau_i#0> with
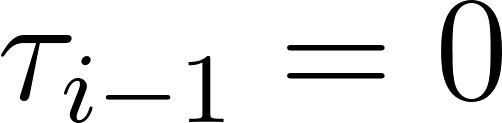
<https://www.codecogs.com/eqnedit.php?latex=%20%5Ctau_%7Bi-1%7D%20%3D%200#0> if
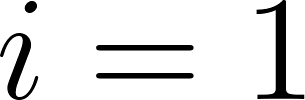
<https://www.codecogs.com/eqnedit.php?latex=i%3D1#0> and
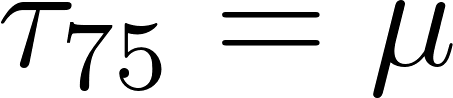
<https://www.codecogs.com/eqnedit.php?latex=%20%5Ctau_%7B75%7D%20%3D%5Cmu#0> as mortality only occurs in the oldest age group, and is equal to the birth rate in the closed system. The scaling of susceptibility dependent on age (including susceptibility reduction due to natural immunity in the first three months) is represented by
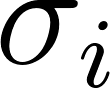
<https://www.codecogs.com/eqnedit.php?latex=%5Csigma_i#0> and
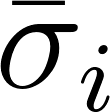
<https://www.codecogs.com/eqnedit.php?latex=%5Cbar%7B%5Csigma%7D_i#0> is the combination of scaling susceptibility due to age and prior exposure. All other parameters are defined in Table S1.

For the immunisation model, variables prefixed by “
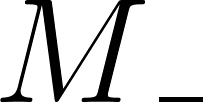
<https://www.codecogs.com/eqnedit.php?latex=M%5C_#0>” indicate individuals are protected by mAb and “
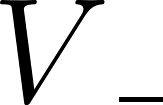
<https://www.codecogs.com/eqnedit.php?latex=V%5C_#0>” indicate individuals are protected by the maternal vaccine. When protected and infected, i.e. in compartments
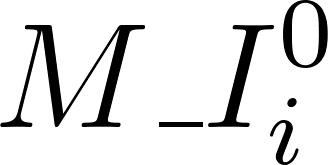
<https://www.codecogs.com/eqnedit.php?latex=M%5C_I%5E0_i#0>,
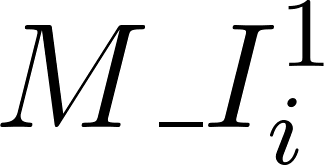
<https://www.codecogs.com/eqnedit.php?latex=M%5C_I%5E1_i#0>or
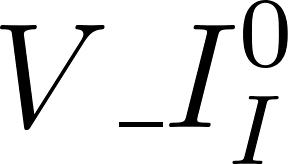
<https://www.codecogs.com/eqnedit.php?latex=V%5C_I%5E0_I#0>, there is no risk of hospitalisation. If
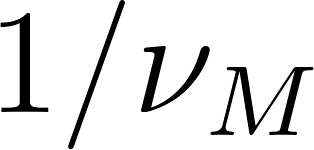
<https://www.codecogs.com/eqnedit.php?latex=1%2F%5Cnu_M#0> is the durability of the mAb, then
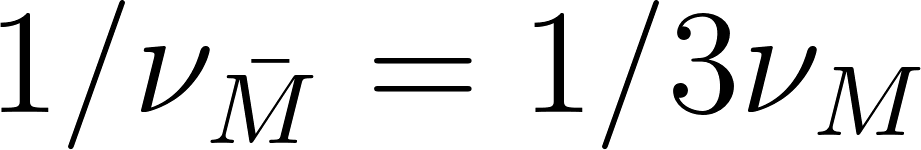
<https://www.codecogs.com/eqnedit.php?latex=1%2F%5Cnu_%7B%5Cbar%7BM%7D%7D%20%3D%201%2F3%5Cnu_M#0> so that on average, an individual is in each of the three mAb protected susceptible compartments for a third of the total duration of protection of the mAb.

In all simulations, the model is “run-in” for a period of 12 years, and then output is checked to ensure solutions have reached equilibrium.

*Base model*

The base model can be represented by the following equations, noting the equations using variables denoted with a bar correspond to the preterm risk group and without the bar, the term population,

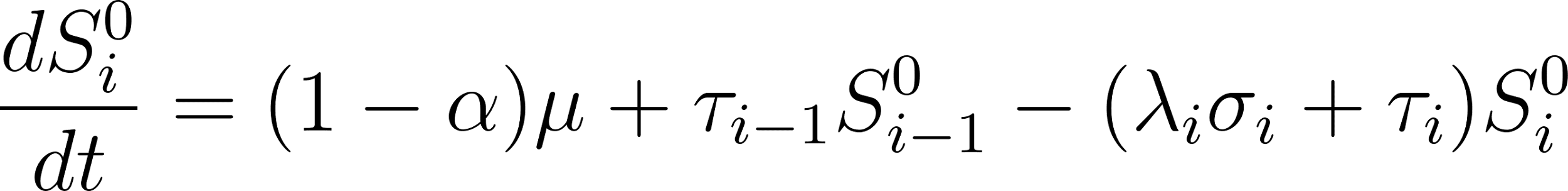
<https://www.codecogs.com/eqnedit.php?latex=%5Cfrac%7Bd%7BS%5E0_i%7D%7D%7Bdt%7D%20%3D%20(1-%20%5Calpha)%5Cmu%20%2B%20%5Ctau_%7Bi-1%7D%7BS%5E0_%7Bi-1%7D%7D%20-%20(%5Clambda_i%20%5Csigma_i%20%2B%20%5Ctau_i)S%5E0_i#0>	(population born at term)

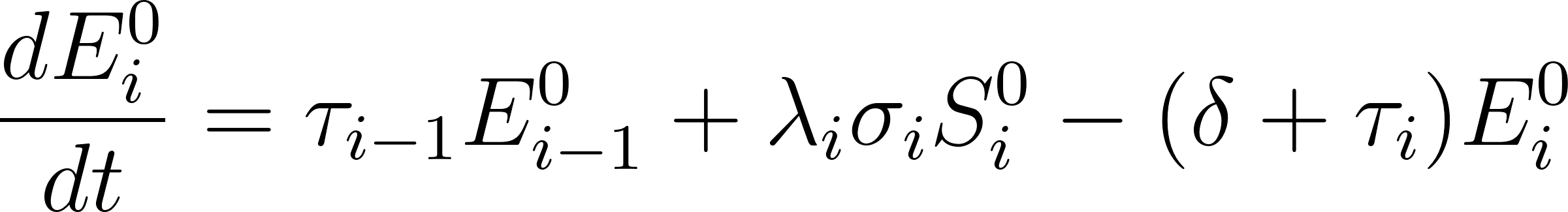
<https://www.codecogs.com/eqnedit.php?latex=%20%5Cfrac%7BdE%5E0_i%7D%7Bdt%7D%20%3D%20%5Ctau_%7Bi-1%7DE%5E0_%7Bi-1%7D%2B%5Clambda_i%20%5Csigma_i%20S%5E0_i%20-%20(%5Cdelta%20%2B%20%5Ctau_i)%20E%5E0_i#0>

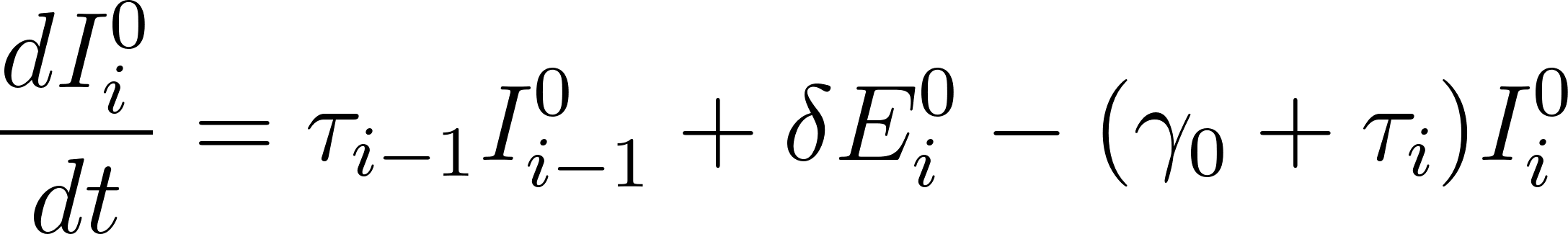
<https://www.codecogs.com/eqnedit.php?latex=%5Cfrac%7BdI%5E0_i%7D%7Bdt%7D%20%3D%20%5Ctau_%7Bi-1%7DI%5E0_%7Bi-1%7D%2B%5Cdelta%20E%5E0_i%20-%20(%5Cgamma_0%20%2B%20%5Ctau_i)%20I%5E0_i#0>

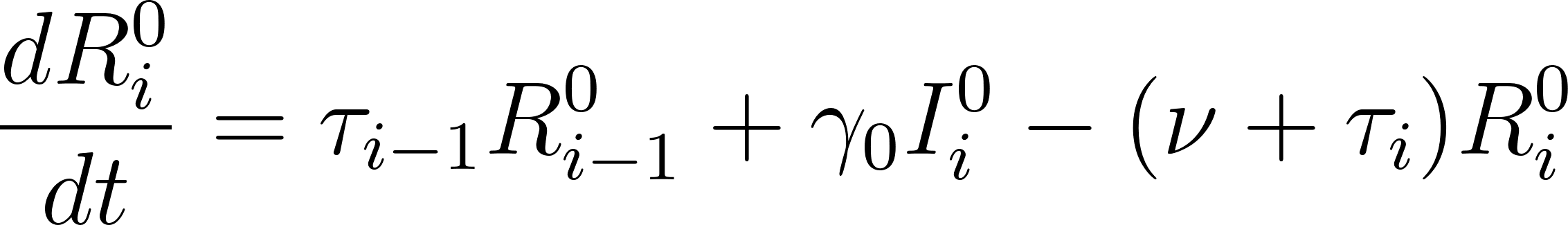
<https://www.codecogs.com/eqnedit.php?latex=%5Cfrac%7BdR%5E0_i%7D%7Bdt%7D%20%3D%20%5Ctau_%7Bi-1%7DR%5E0_%7Bi-1%7D%2B%5Cgamma_0%20I%5E0_i%20-%20(%5Cnu%20%2B%20%5Ctau_i)%20R%5E0_i#0>

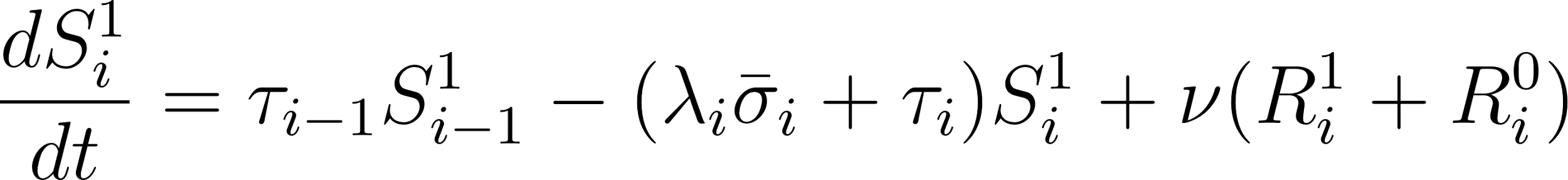
<https://www.codecogs.com/eqnedit.php?latex=%5Cfrac%7BdS%5E1_i%7D%7Bdt%7D%20%3D%20%5Ctau_%7Bi-1%7DS%5E1_%7Bi-1%7D%20-%20(%5Clambda_i%20%5Cbar%7B%5Csigma%7D_i%20%2B%20%5Ctau_i)S%5E1_i%20%2B%20%5Cnu%20(R%5E1_i%20%2B%20R%5E0_i)#0>

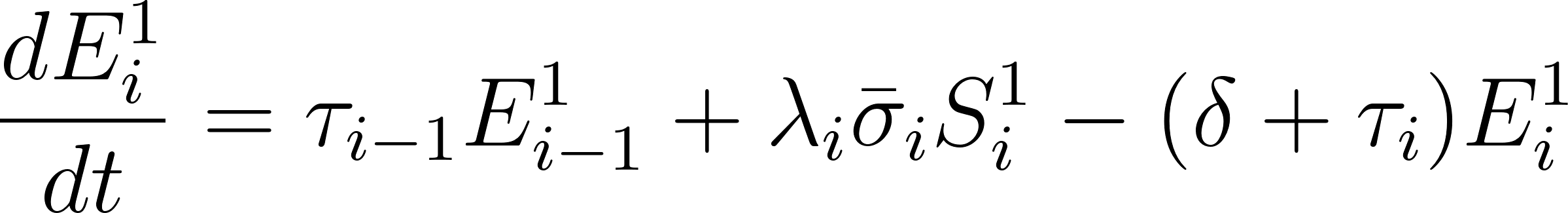
<https://www.codecogs.com/eqnedit.php?latex=%20%5Cfrac%7BdE%5E1_i%7D%7Bdt%7D%20%3D%20%5Ctau_%7Bi-1%7DE%5E1_%7Bi-1%7D%2B%5Clambda_i%20%5Cbar%7B%5Csigma%7D_i%20S%5E1_i%20-%20(%5Cdelta%20%2B%20%5Ctau_i)%20E%5E1_i#0>

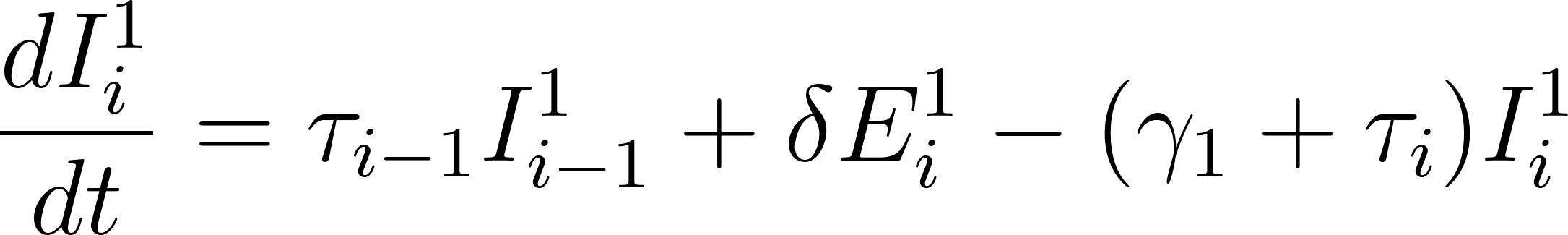
<https://www.codecogs.com/eqnedit.php?latex=%5Cfrac%7BdI%5E1_i%7D%7Bdt%7D%20%3D%20%5Ctau_%7Bi-1%7DI%5E1_%7Bi-1%7D%2B%5Cdelta%20E%5E1_i%20-%20(%5Cgamma_1%20%2B%20%5Ctau_i)%20I%5E1_i#0>

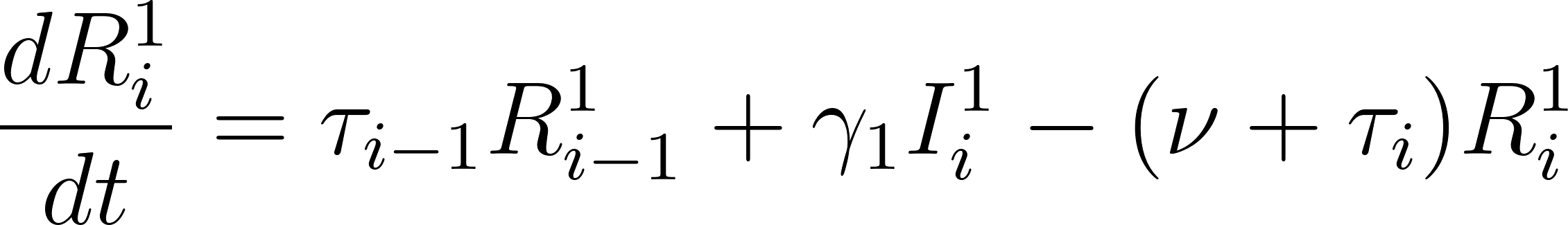
<https://www.codecogs.com/eqnedit.php?latex=%5Cfrac%7BdR%5E1_i%7D%7Bdt%7D%20%3D%20%5Ctau_%7Bi-1%7DR%5E1_%7Bi-1%7D%2B%5Cgamma_1%20I%5E1_i%20-%20(%5Cnu%20%2B%20%5Ctau_i)%20R%5E1_i#0>

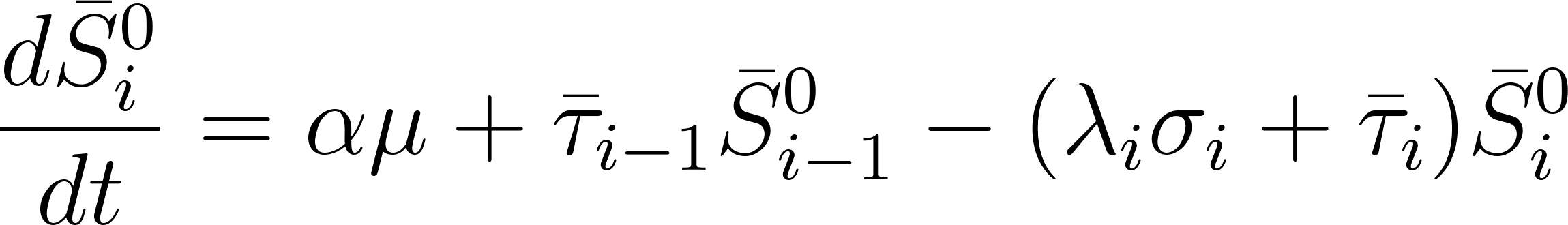
<https://www.codecogs.com/eqnedit.php?latex=%5Cfrac%7Bd%7B%5Cbar%7BS%7D%5E0_i%7D%7D%7Bdt%7D%20%3D%20%5Calpha%5Cmu%20%2B%20%5Cbar%7B%5Ctau%7D_%7Bi-1%7D%7B%5Cbar%7BS%7D%5E0_%7Bi-1%7D%7D%20-%20(%5Clambda_i%20%5Csigma_i%20%2B%20%5Cbar%7B%5Ctau%7D_i)%5Cbar%7BS%7D%5E0_i#0>	(population born preterm)

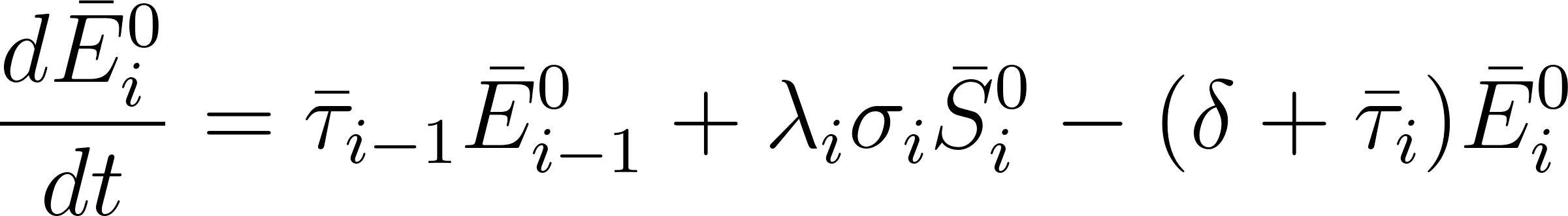
<https://www.codecogs.com/eqnedit.php?latex=%20%5Cfrac%7Bd%5Cbar%7BE%7D%5E0_i%7D%7Bdt%7D%20%3D%20%5Cbar%7B%5Ctau%7D_%7Bi-1%7D%5Cbar%7BE%7D%5E0_%7Bi-1%7D%2B%5Clambda_i%20%5Csigma_i%20%5Cbar%7BS%7D%5E0_i%20-%20(%5Cdelta%20%2B%20%20%5Cbar%7B%5Ctau%7D_i)%20%5Cbar%7BE%7D%5E0_i#0>

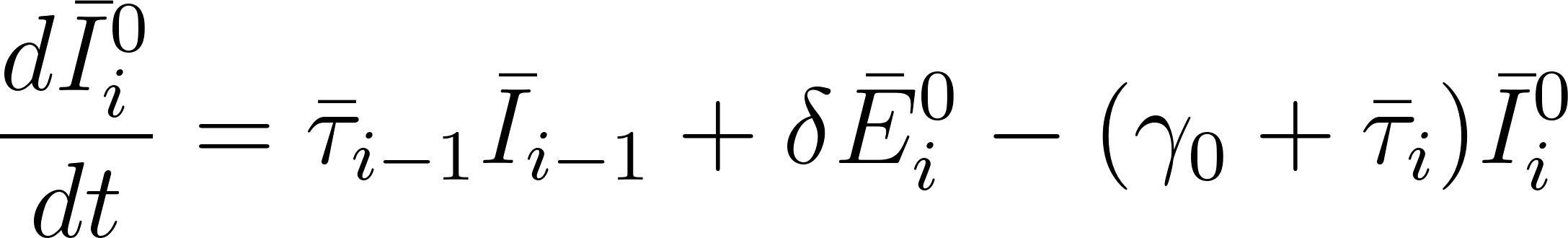
<https://www.codecogs.com/eqnedit.php?latex=%5Cfrac%7Bd%5Cbar%7BI%7D%5E0_i%7D%7Bdt%7D%20%3D%20%20%5Cbar%7B%5Ctau%7D_%7Bi-1%7D%5Cbar%7BI%7D_%7Bi-1%7D%2B%5Cdelta%20%5Cbar%7BE%7D%5E0_i%20-%20(%5Cgamma_0%20%2B%20%20%5Cbar%7B%5Ctau%7D_i)%20%5Cbar%7BI%7D%5E0_i#0>

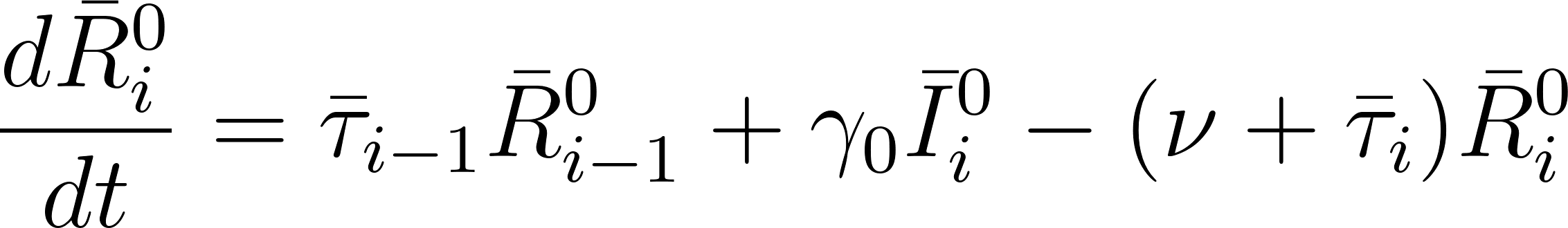
<https://www.codecogs.com/eqnedit.php?latex=%5Cfrac%7Bd%5Cbar%7BR%7D%5E0_i%7D%7Bdt%7D%20%3D%20%20%5Cbar%7B%5Ctau%7D_%7Bi-1%7D%5Cbar%7BR%7D%5E0_%7Bi-1%7D%2B%5Cgamma_0%20%5Cbar%7BI%7D%5E0_i%20-%20(%5Cnu%20%2B%20%20%5Cbar%7B%5Ctau%7D_i)%20%5Cbar%7BR%7D%5E0_i#0>

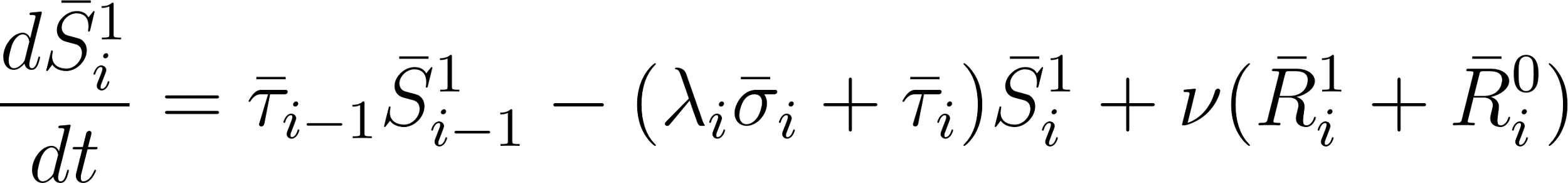
<https://www.codecogs.com/eqnedit.php?latex=%5Cfrac%7Bd%5Cbar%7BS%7D%5E1_i%7D%7Bdt%7D%20%3D%20%20%5Cbar%7B%5Ctau%7D_%7Bi-1%7D%5Cbar%7BS%7D%5E1_%7Bi-1%7D%20-%20(%5Clambda_i%20%5Cbar%7B%5Csigma%7D_i%20%2B%20%20%5Cbar%7B%5Ctau%7D_i)%5Cbar%7BS%7D%5E1_i%20%2B%20%5Cnu%20(%5Cbar%7BR%7D%5E1_i%20%2B%20%5Cbar%7BR%7D%5E0_i)#0>

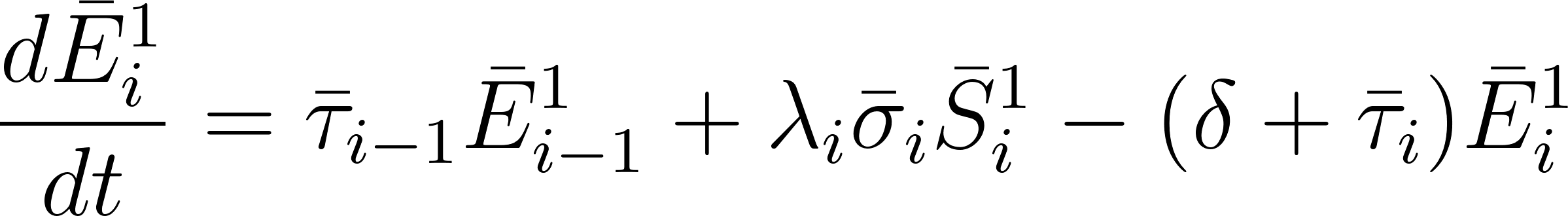
<https://www.codecogs.com/eqnedit.php?latex=%20%5Cfrac%7Bd%5Cbar%7BE%7D%5E1_i%7D%7Bdt%7D%20%3D%20%20%5Cbar%7B%5Ctau%7D_%7Bi-1%7D%5Cbar%7BE%7D%5E1_%7Bi-1%7D%2B%5Clambda_i%20%5Cbar%7B%5Csigma%7D_i%20%5Cbar%7BS%7D%5E1_i%20-%20(%5Cdelta%20%2B%20%20%5Cbar%7B%5Ctau%7D_i)%20%5Cbar%7BE%7D%5E1_i#0>

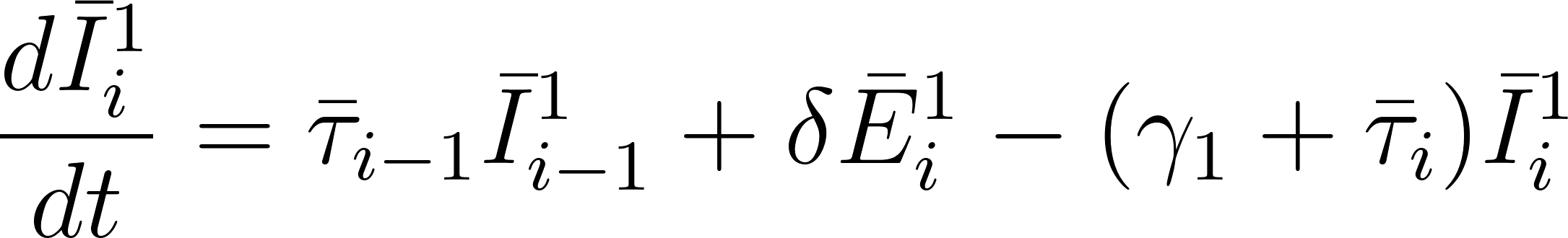
<https://www.codecogs.com/eqnedit.php?latex=%5Cfrac%7Bd%5Cbar%7BI%7D%5E1_i%7D%7Bdt%7D%20%3D%20%20%5Cbar%7B%5Ctau%7D_%7Bi-1%7D%5Cbar%7BI%7D%5E1_%7Bi-1%7D%2B%5Cdelta%20%5Cbar%7BE%7D%5E1_i%20-%20(%5Cgamma_1%20%2B%20%20%5Cbar%7B%5Ctau%7D_i)%20%5Cbar%7BI%7D%5E1_i#0>

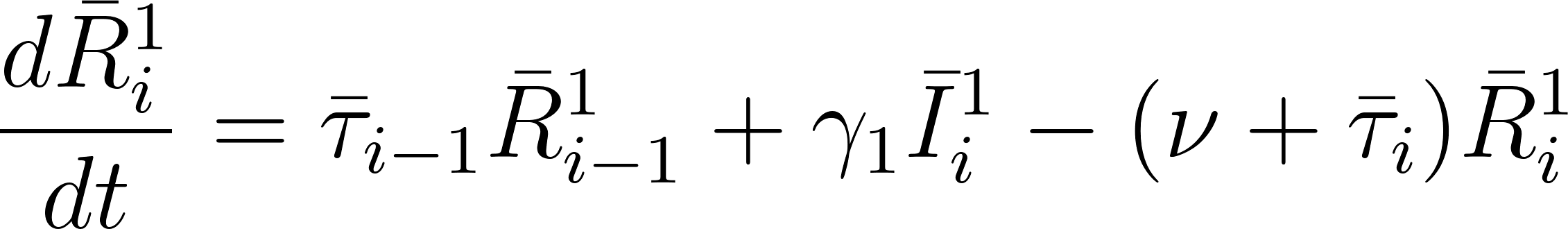
<https://www.codecogs.com/eqnedit.php?latex=%5Cfrac%7Bd%5Cbar%7BR%7D%5E1_i%7D%7Bdt%7D%20%3D%20%20%5Cbar%7B%5Ctau%7D_%7Bi-1%7D%5Cbar%7BR%7D%5E1_%7Bi-1%7D%2B%5Cgamma_1%20%5Cbar%7BI%7D%5E1_i%20-%20(%5Cnu%20%2B%20%20%5Cbar%7B%5Ctau%7D_i)%20%5Cbar%7BR%7D%5E1_i#0>

The transmission function is represented by the following,

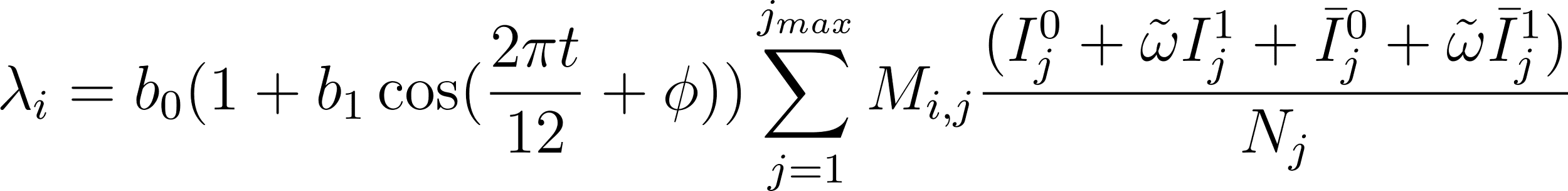
<https://www.codecogs.com/eqnedit.php?latex=%5Clambda_i%20%3D%20b_0(1%2Bb_1%5Ccos%20(%5Cfrac%7B2%5Cpi%20t%7D%7B12%7D%2B%5Cphi))%5Csum_%7Bj%3D1%7D%5E%7Bj_%7Bmax%7D%7DM_%7Bi%2Cj%7D%5Cfrac%7B(I%5E0_j%2B%5Ctilde%7B%5Comega%7DI%5E1_j%2B%20%5Cbar%7BI%7D%5E0_j%2B%5Ctilde%7B%5Comega%7D%5Cbar%7BI%7D%5E1_j)%7D%7BN_j%7D#0>

*Immunisation model*

We show here the equations for the immunisation model, combining the term and preterm populations for clarity. These equations are extended in the same way as shown in the base model equations, to stratify the term and preterm populations.

(unprotected)

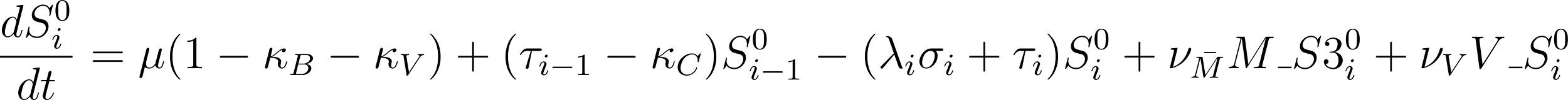
<https://www.codecogs.com/eqnedit.php?latex=%5Cfrac%7Bd%7BS%5E0_i%7D%7D%7Bdt%7D%20%3D%20%5Cmu%20(1-%5Ckappa_B-%5Ckappa_V)%2B%20(%5Ctau_%7Bi-1%7D-%5Ckappa_C)%7BS%5E0_%7Bi-1%7D%7D%20-%20(%5Clambda_i%20%5Csigma_i%20%2B%20%5Ctau_i)S%5E0_i%20%2B%20%5Cnu_%7B%5Cbar%7BM%7D%7D%20M%5C_S3%5E0_i%20%2B%20%5Cnu_V%20V%5C_S%5E0_i#0>

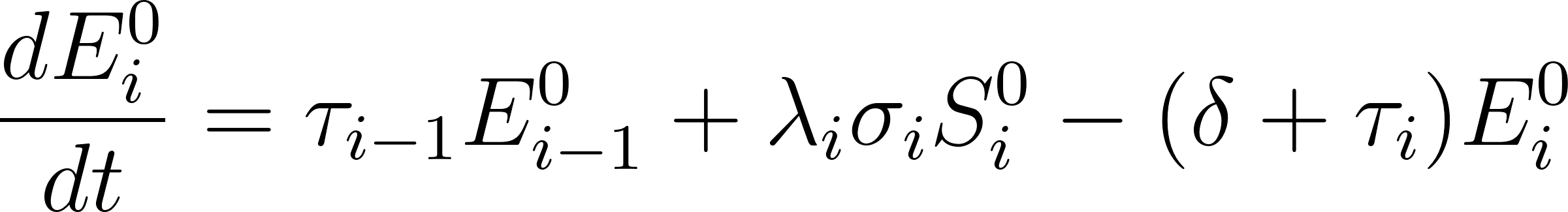
<https://www.codecogs.com/eqnedit.php?latex=%20%5Cfrac%7BdE%5E0_i%7D%7Bdt%7D%20%3D%20%5Ctau_%7Bi-1%7DE%5E0_%7Bi-1%7D%2B%5Clambda_i%20%5Csigma_i%20S%5E0_i%20-%20(%5Cdelta%20%2B%20%5Ctau_i)%20E%5E0_i#0>

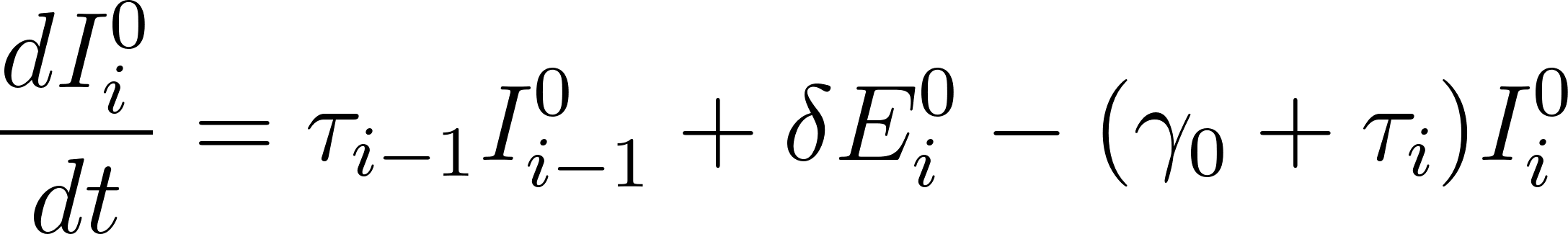
<https://www.codecogs.com/eqnedit.php?latex=%5Cfrac%7BdI%5E0_i%7D%7Bdt%7D%20%3D%20%5Ctau_%7Bi-1%7DI%5E0_%7Bi-1%7D%2B%5Cdelta%20E%5E0_i%20-%20(%5Cgamma_0%20%2B%20%5Ctau_i)%20I%5E0_i#0>

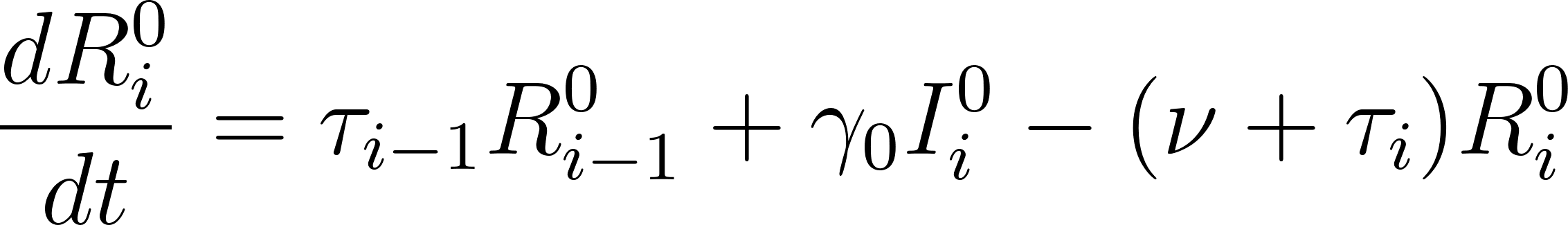
<https://www.codecogs.com/eqnedit.php?latex=%5Cfrac%7BdR%5E0_i%7D%7Bdt%7D%20%3D%20%5Ctau_%7Bi-1%7DR%5E0_%7Bi-1%7D%2B%5Cgamma_0%20I%5E0_i%20-%20(%5Cnu%20%2B%20%5Ctau_i)%20R%5E0_i#0>

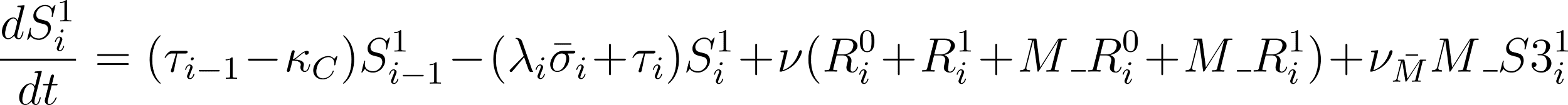
<https://www.codecogs.com/eqnedit.php?latex=%5Cfrac%7BdS%5E1_i%7D%7Bdt%7D%20%3D%20(%5Ctau_%7Bi-1%7D-%5Ckappa_C)S%5E1_%7Bi-1%7D%20-%20(%5Clambda_i%20%5Cbar%7B%5Csigma%7D_i%20%2B%20%5Ctau_i)S%5E1_i%20%2B%20%5Cnu%20(R%5E0_i%20%2B%20R%5E1_i%20%2B%20M%5C_R%5E0_i%20%2B%20M%5C_R%5E1_i)%20%2B%20%5Cnu_%7B%5Cbar%7BM%7D%7D%20M%5C_S3%5E1_i#0>

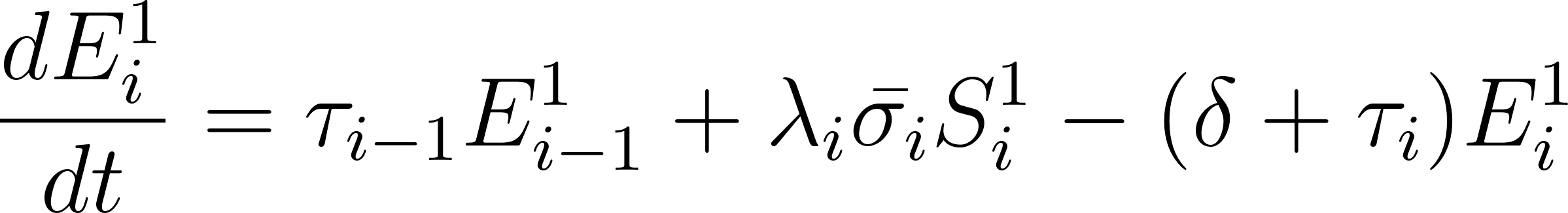
<https://www.codecogs.com/eqnedit.php?latex=%20%5Cfrac%7BdE%5E1_i%7D%7Bdt%7D%20%3D%20%5Ctau_%7Bi-1%7DE%5E1_%7Bi-1%7D%2B%5Clambda_i%20%5Cbar%7B%5Csigma_i%7D%20S%5E1_i%20-%20(%5Cdelta%20%2B%20%5Ctau_i)%20E%5E1_i#0>

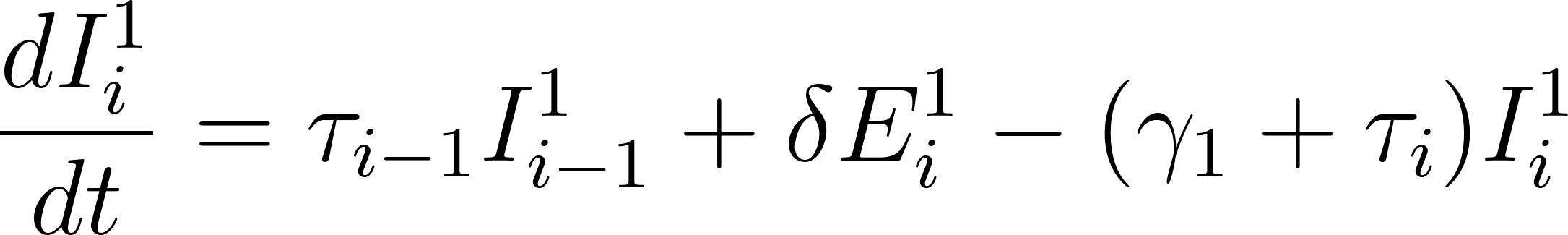
<https://www.codecogs.com/eqnedit.php?latex=%5Cfrac%7BdI%5E1_i%7D%7Bdt%7D%20%3D%20%5Ctau_%7Bi-1%7DI%5E1_%7Bi-1%7D%2B%5Cdelta%20E%5E1_i%20-%20(%5Cgamma_1%20%2B%20%5Ctau_i)%20I%5E1_i#0>

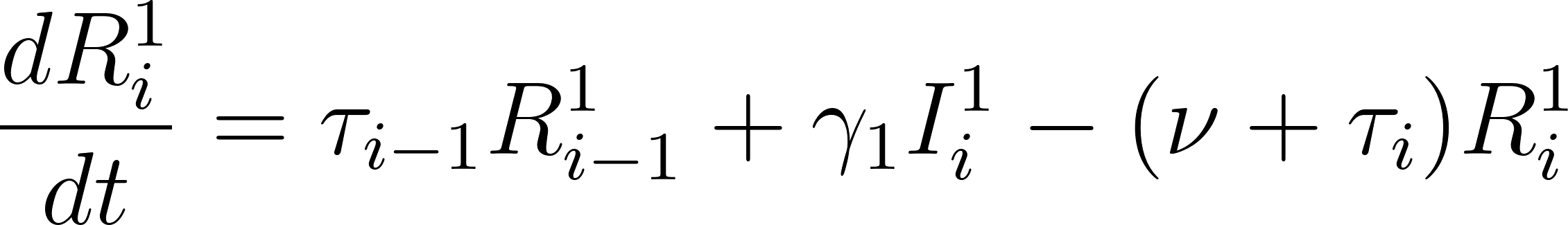
<https://www.codecogs.com/eqnedit.php?latex=%5Cfrac%7BdR%5E1_i%7D%7Bdt%7D%20%3D%20%5Ctau_%7Bi-1%7DR%5E1_%7Bi-1%7D%2B%5Cgamma_1%20I%5E1_i%20-%20(%5Cnu%20%2B%20%5Ctau_i)%20R%5E1_i#0>

(protected maternal vaccine)

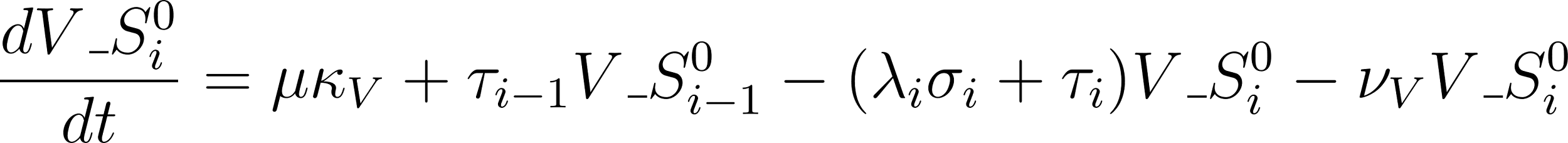
<https://www.codecogs.com/eqnedit.php?latex=%5Cfrac%7BdV%5C_S%5E0_i%7D%7Bdt%7D%20%3D%20%5Cmu%5Ckappa_V%20%20%2B%20%5Ctau_%7Bi-1%7DV%5C_S%5E0_%7Bi-1%7D%20-%20(%5Clambda_i%20%5Csigma_i%20%2B%20%5Ctau_i)V%5C_S%5E0_i%20-%20%5Cnu_V%20%20V%5C_S%5E0_i#0>

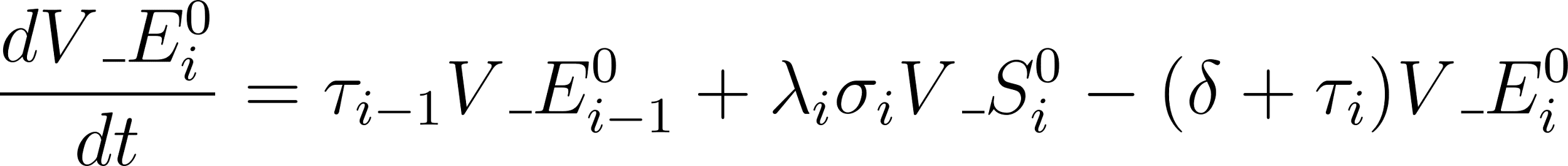
<https://www.codecogs.com/eqnedit.php?latex=%20%5Cfrac%7BdV%5C_E%5E0_i%7D%7Bdt%7D%20%3D%20%5Ctau_%7Bi-1%7DV%5C_E%5E0_%7Bi-1%7D%2B%5Clambda_i%20%5Csigma_i%20V%5C_S%5E0_i%20-%20(%5Cdelta%20%2B%20%5Ctau_i)%20V%5C_E%5E0_i#0>

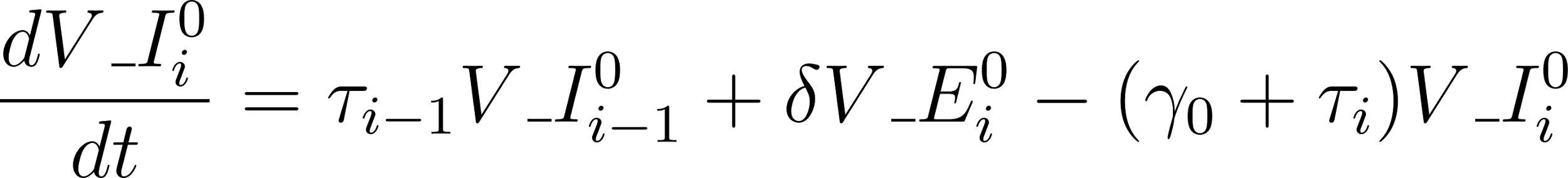
<https://www.codecogs.com/eqnedit.php?latex=%5Cfrac%7BdV%5C_I%5E0_i%7D%7Bdt%7D%20%3D%20%5Ctau_%7Bi-1%7DV%5C_I%5E0_%7Bi-1%7D%2B%5Cdelta%20V%5C_E%5E0_i%20-%20(%5Cgamma_0%20%2B%20%5Ctau_i)%20V%5C_I%5E0_i#0>

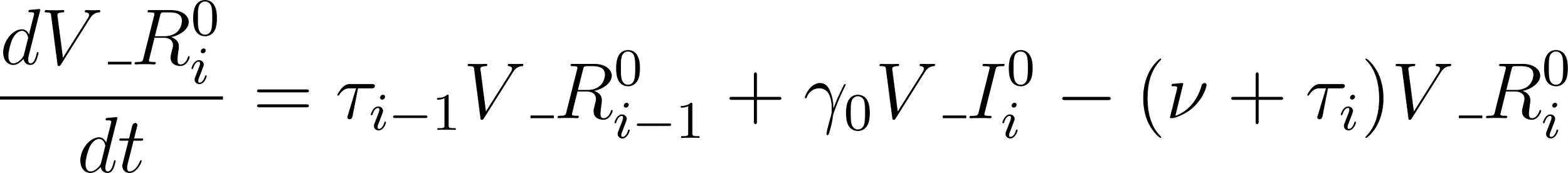
<https://www.codecogs.com/eqnedit.php?latex=%5Cfrac%7BdV%5C_R%5E0_i%7D%7Bdt%7D%20%3D%20%5Ctau_%7Bi-1%7DV%5C_R%5E0_%7Bi-1%7D%2B%5Cgamma_0%20V%5C_I%5E0_i%20-%20(%5Cnu%20%2B%20%5Ctau_i)%20V%5C_R%5E0_i#0>

(protected mAb)

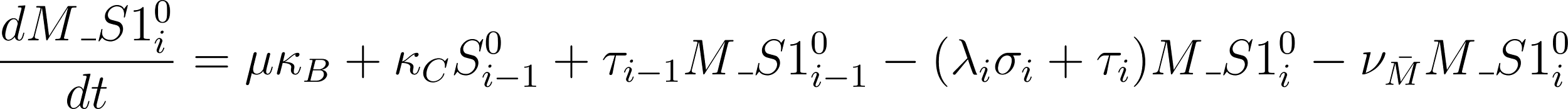
<https://www.codecogs.com/eqnedit.php?latex=%5Cfrac%7Bd%7BM%5C_S1%5E0_i%7D%7D%7Bdt%7D%20%3D%20%5Cmu%5Ckappa_B%20%2B%20%5Ckappa_C%20S%5E0_%7Bi-1%7D%2B%20%5Ctau_%7Bi-1%7D%7BM%5C_S1%5E0_%7Bi-1%7D%7D%20-%20(%5Clambda_i%20%5Csigma_i%20%2B%20%5Ctau_i)M%5C_S1%5E0_i%20-%20%5Cnu_%7B%5Cbar%7BM%7D%7D%20M%5C_S1%5E0_i#0>

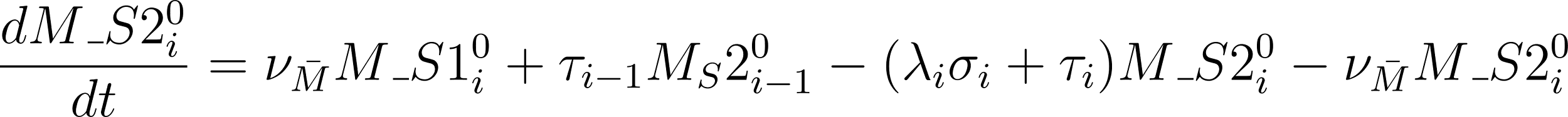
<https://www.codecogs.com/eqnedit.php?latex=%5Cfrac%7Bd%7BM%5C_S2%5E0_i%7D%7D%7Bdt%7D%20%3D%20%5Cnu_%7B%5Cbar%7BM%7D%7D%20M%5C_S1%5E0_i%20%2B%20%5Ctau_%7Bi-1%7D%7BM_S2%5E0_%7Bi-1%7D%7D%20-%20(%5Clambda_i%20%5Csigma_i%20%2B%20%5Ctau_i)M%5C_S2%5E0_i%20-%20%5Cnu_%7B%5Cbar%7BM%7D%7D%20%20M%5C_S2%5E0_i#0>

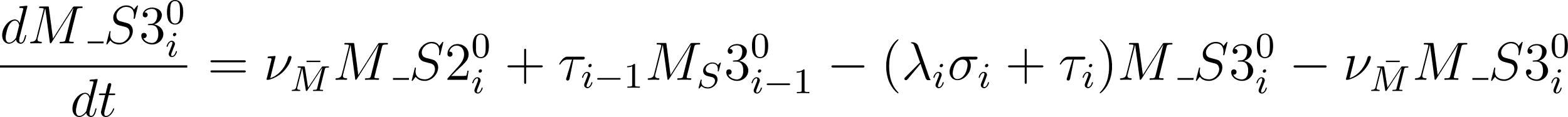
<https://www.codecogs.com/eqnedit.php?latex=%5Cfrac%7Bd%7BM%5C_S3%5E0_i%7D%7D%7Bdt%7D%20%3D%5Cnu_%7B%5Cbar%7BM%7D%7D%20%20M%5C_S2%5E0_i%20%2B%20%5Ctau_%7Bi-1%7D%7BM_S3%5E0_%7Bi-1%7D%7D%20-%20(%5Clambda_i%20%5Csigma_i%20%2B%20%5Ctau_i)M%5C_S3%5E0_i%20-%20%5Cnu_%7B%5Cbar%7BM%7D%7D%20%20M%5C_S3%5E0_i#0>

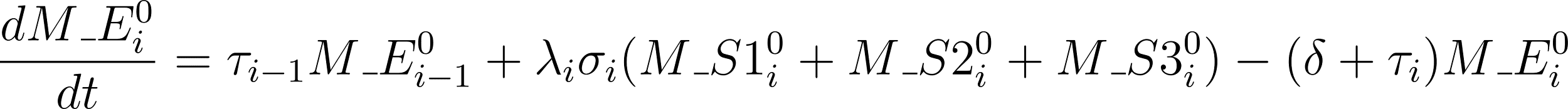
<https://www.codecogs.com/eqnedit.php?latex=%20%5Cfrac%7BdM%5C_E%5E0_i%7D%7Bdt%7D%20%3D%20%5Ctau_%7Bi-1%7DM%5C_E%5E0_%7Bi-1%7D%2B%5Clambda_i%20%5Csigma_i%20(M%5C_S1%5E0_i%20%2B%20M%5C_S2%5E0_i%20%2B%20M%5C_S3%5E0_i)%20-%20(%5Cdelta%20%2B%20%5Ctau_i)%20M%5C_E%5E0_i#0>

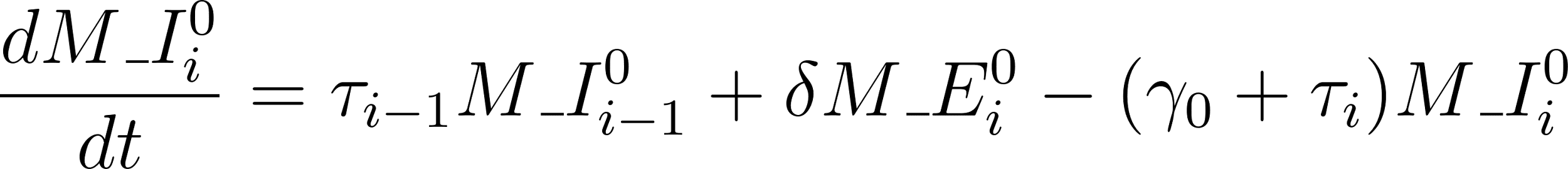
<https://www.codecogs.com/eqnedit.php?latex=%5Cfrac%7BdM%5C_I%5E0_i%7D%7Bdt%7D%20%3D%20%5Ctau_%7Bi-1%7DM%5C_I%5E0_%7Bi-1%7D%2B%5Cdelta%20M%5C_E%5E0_i%20-%20(%5Cgamma_0%20%2B%20%5Ctau_i)%20M%5C_I%5E0_i#0>

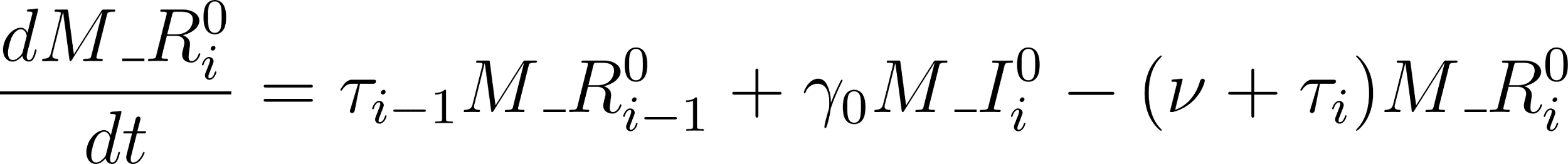
<https://www.codecogs.com/eqnedit.php?latex=%5Cfrac%7BdM%5C_R%5E0_i%7D%7Bdt%7D%20%3D%20%5Ctau_%7Bi-1%7DM%5C_R%5E0_%7Bi-1%7D%2B%5Cgamma_0%20M%5C_I%5E0_i%20-%20(%5Cnu%20%2B%20%5Ctau_i)%20M%5C_R%5E0_i#0>

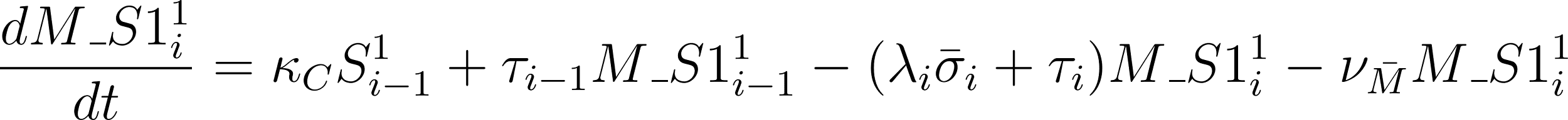
<https://www.codecogs.com/eqnedit.php?latex=%5Cfrac%7BdM%5C_S1%5E1_i%7D%7Bdt%7D%20%3D%20%5Ckappa__C%20S%5E1_%7Bi-1%7D%20%2B%20%5Ctau_%7Bi-1%7DM%5C_S1%5E1_%7Bi-1%7D%20-%20(%5Clambda_i%20%5Cbar%7B%5Csigma%7D_i%20%2B%20%5Ctau_i)M%5C_S1%5E1_i%20-%20%5Cnu_%7B%5Cbar%7BM%7D%7D%20%20M%5C_S1%5E1_i#0>

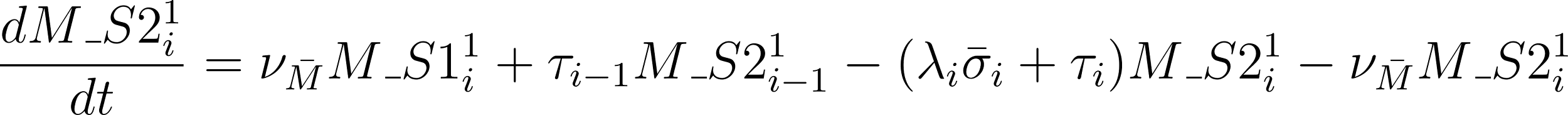
<https://www.codecogs.com/eqnedit.php?latex=%5Cfrac%7BdM%5C_S2%5E1_i%7D%7Bdt%7D%20%3D%20%5Cnu_%7B%5Cbar%7BM%7D%7D%20%20M%5C_S1%5E1_i%20%2B%20%5Ctau_%7Bi-1%7DM%5C_S2%5E1_%7Bi-1%7D%20-%20(%5Clambda_i%20%5Cbar%7B%5Csigma%7D_i%20%2B%20%5Ctau_i)M%5C_S2%5E1_i%20-%5Cnu_%7B%5Cbar%7BM%7D%7D%20%20M%5C_S2%5E1_i#0>

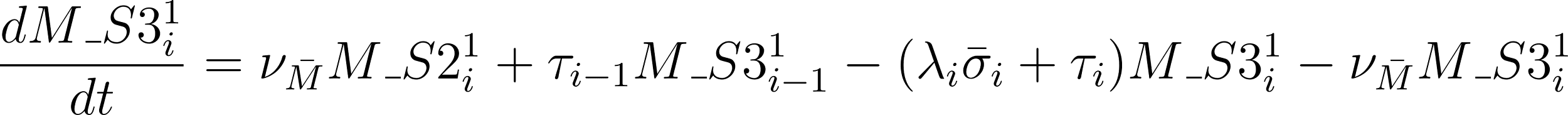
<https://www.codecogs.com/eqnedit.php?latex=%5Cfrac%7BdM%5C_S3%5E1_i%7D%7Bdt%7D%20%3D%20%5Cnu_%7B%5Cbar%7BM%7D%7D%20%20M%5C_S2%5E1_i%20%2B%20%5Ctau_%7Bi-1%7DM%5C_S3%5E1_%7Bi-1%7D%20-%20(%5Clambda_i%20%5Cbar%7B%5Csigma%7D_i%20%2B%20%5Ctau_i)M%5C_S3%5E1_i%20-%20%5Cnu_%7B%5Cbar%7BM%7D%7D%20%20M%5C_S3%5E1_i#0>

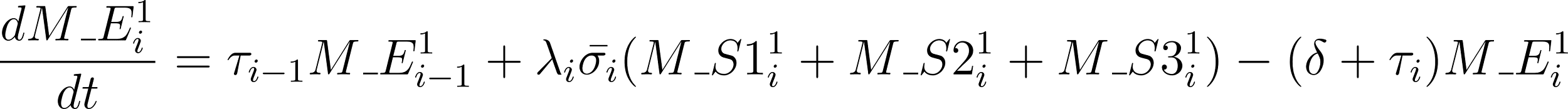
<https://www.codecogs.com/eqnedit.php?latex=%20%5Cfrac%7BdM%5C_E%5E1_i%7D%7Bdt%7D%20%3D%20%5Ctau_%7Bi-1%7DM%5C_E%5E1_%7Bi-1%7D%2B%5Clambda_i%20%5Cbar%7B%5Csigma_i%7D%20(M%5C_S1%5E1_i%20%2B%20M%5C_S2%5E1_i%20%2B%20M%5C_S3%5E1_i)%20-%20(%5Cdelta%20%2B%20%5Ctau_i)%20M%5C_E%5E1_i#0>

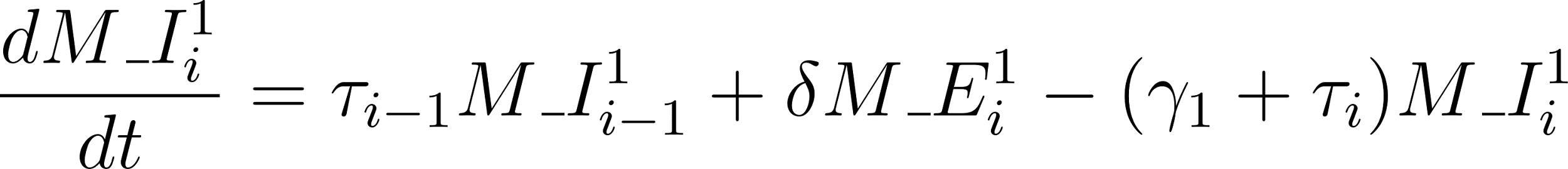
<https://www.codecogs.com/eqnedit.php?latex=%5Cfrac%7BdM%5C_I%5E1_i%7D%7Bdt%7D%20%3D%20%5Ctau_%7Bi-1%7DM%5C_I%5E1_%7Bi-1%7D%2B%5Cdelta%20M%5C_E%5E1_i%20-%20(%5Cgamma_1%20%2B%20%5Ctau_i)%20M%5C_I%5E1_i#0>

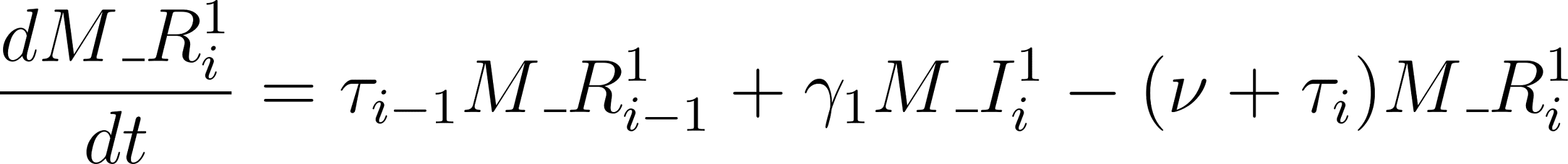
<https://www.codecogs.com/eqnedit.php?latex=%5Cfrac%7BdM%5C_R%5E1_i%7D%7Bdt%7D%20%3D%20%5Ctau_%7Bi-1%7DM%5C_R%5E1_%7Bi-1%7D%2B%5Cgamma_1%20M%5C_I%5E1_i%20-%20(%5Cnu%20%2B%20%5Ctau_i)%20M%5C_R%5E1_i#0>

The transmission function is represented by the following,

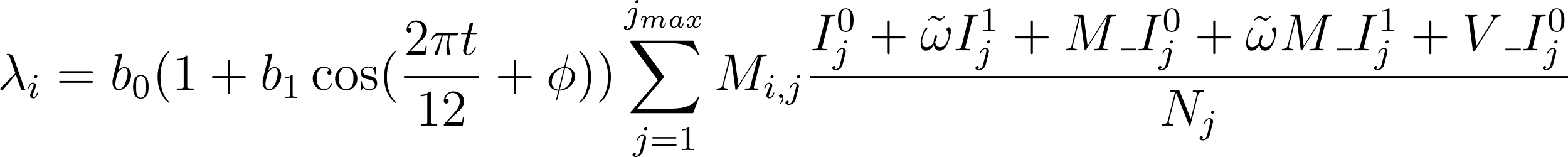
<https://www.codecogs.com/eqnedit.php?latex=%5Clambda_i%20%3D%20b_0(1%2Bb_1%5Ccos%20(%5Cfrac%7B2%5Cpi%20t%7D%7B12%7D%2B%5Cphi))%5Csum_%7Bj%3D1%7D%5E%7Bj_%7Bmax%7D%7DM_%7Bi%2Cj%7D%20%5Cfrac%7BI%5E0_j%2B%5Ctilde%7B%5Comega%7DI%5E1_j%2B%20M%5C_I%5E0_j%2B%5Ctilde%7B%5Comega%7DM%5C_I%5E1_j%2BV%5C_I%5E0_j%7D%7BN_j%7D#0>

### Contact matrix

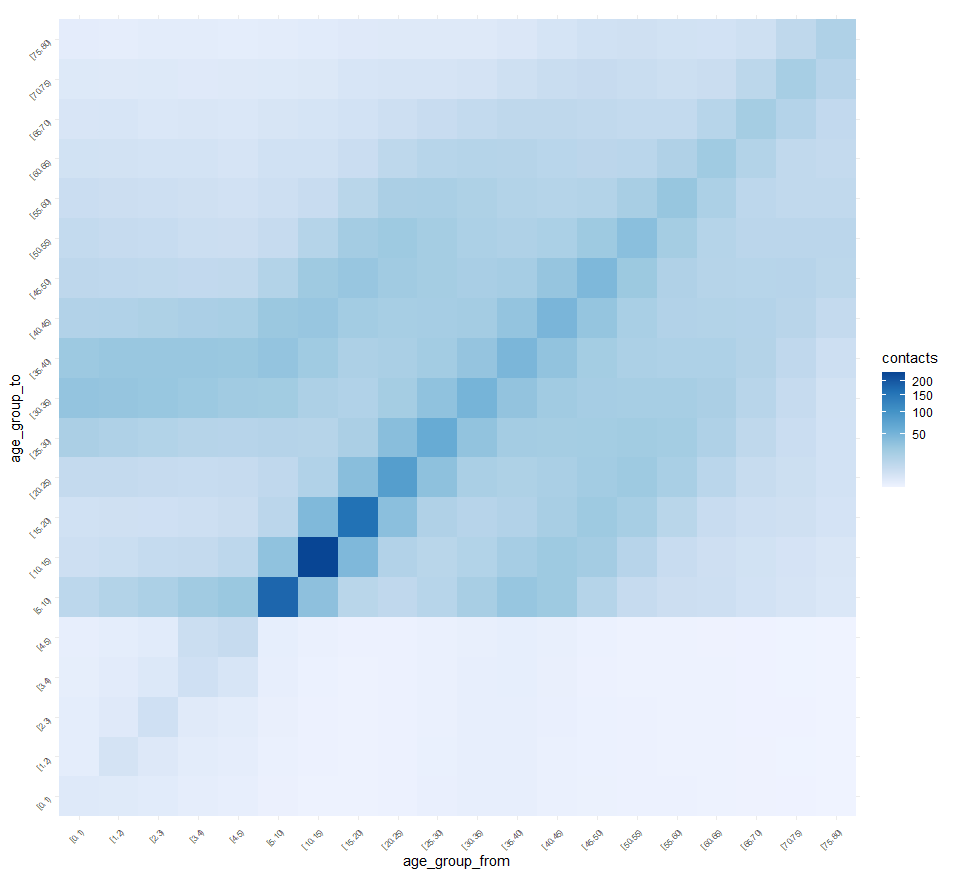

**Figure S2**: Monthly number of contacts between five-year age groups for ages >= 5 years old, and yearly for those < 5 years old. This contact matrix was generated using the R package *conmat* based on the POLYMOD study results for the UK, with household contacts based on the demographics of the 2016 ABS 5% microdata sample for the Greater Perth region and between school/childcare contacts of children under 5 based on the Childhood Education and Care Survey, normalised to 2016 southern Western Australia population demographics. The yearly age groups < 5 are divided into monthly age groups in the model, with contacts being uniformly distributed into these resulting age groups.

### Model fitting

We used a Markov-chain Monte Carlo (MCMC) process with the Metropolis-Hastings algorithm for maximum likelihood estimation as implemented in the R package *lazymcmc* to fit the model to monthly RSV-hospitalisations from five age groups (0- 2 months, 3 - 5 months, 6 - 11 months, 12 - 23 months and 2 - <5 years), stratified by term and preterm (see Figure S1) to fit the scaling of increased risk of hospitalisation due to RSV infection for preterm infants as compared to term. To calibrate the level of infection in the model, the fitting also captured the criteria that most children have experienced an RSV infection by the age of two years, as described below [[7,19]](https://paperpile.com/c/Y68o4Y/oZ1I0+b1QRn).

#### Age-to-risk function

We translated modelled infections to hospitalisations using a function relating age at acquisition of infection to the risk of hospitalisation. The function scales baseline risk in individuals with no prior infection to the (lower) risk in individuals with at least one prior infection, and preterm (higher) risk to baseline term risk. We assumed the age-to-risk function takes an exponential decay form such that

$y=(Ae^{-Bt}+C)DE$,

where *y* is the modelled probabilistic risk of hospitalisation at age *t* (in months) given an individual is infected with RSV, with *A* the average maximum risk increase over the minimum C for a term infant (at age 0), B the exponential function decay constant, D a scaling factor (<1) modifying risk for the prior infection group reflecting a lower risk of hospitalisation, and E a scaling factor (>1) modifying risk for those born preterm reflecting a higher risk of hospitalisation. We estimate A, B, C and E through model fitting.

#### Log likelihood

We fit the parameters b_0,_ b_1,_ A, B, C and E to RSV-hospitalisation data using MCMC for maximum likelihood estimation. To derive the log likelihood, we assumed that the number of hospitalisations each month followed a Poisson distribution. For the model with the term/preterm stratification, the log likelihood,
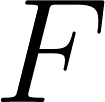
<https://www.codecogs.com/eqnedit.php?latex=F#0> is the sum of the log likelihood of the term,
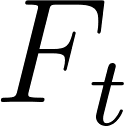
<https://www.codecogs.com/eqnedit.php?latex=F_t#0>, and preterm,
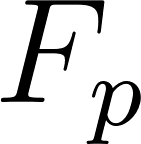
<https://www.codecogs.com/eqnedit.php?latex=F_p#0>, components of the model,

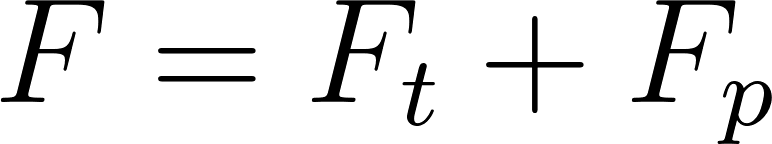
<https://www.codecogs.com/eqnedit.php?latex=F%20%3D%20F_t%20%2B%20F_p#0>

with

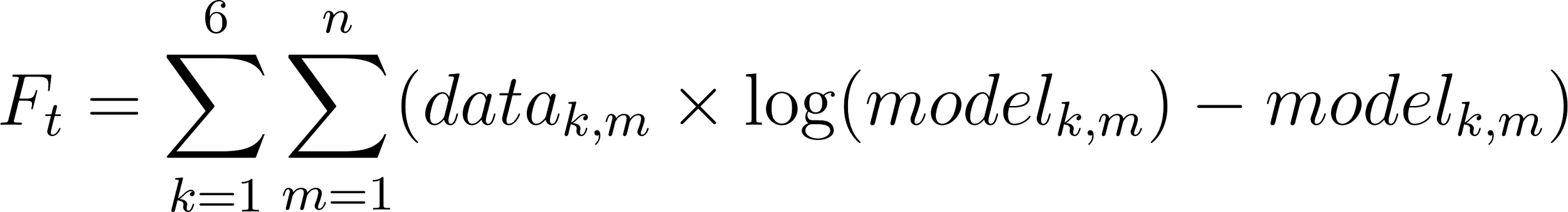
<https://www.codecogs.com/eqnedit.php?latex=F_t%20%3D%20%5Csum_%7Bk%3D1%7D%5E%7B6%7D%5Csum_%7Bm%3D1%7D%5E%7Bn%7D(data_%7Bk%2Cm%7D%20%5Ctimes%20%5Clog(model_%7Bk%2Cm%7D)%20-%20model_%7Bk%2Cm%7D)#0>

and

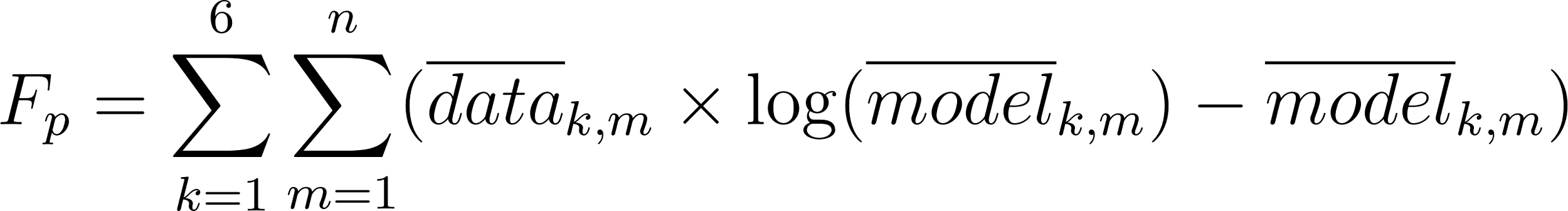
<https://www.codecogs.com/eqnedit.php?latex=F_p%20%3D%20%5Csum_%7Bk%3D1%7D%5E%7B6%7D%5Csum_%7Bm%3D1%7D%5E%7Bn%7D(%5Coverline%7Bdata%7D_%7Bk%2Cm%7D%20%5Ctimes%20%5Clog(%5Coverline%7Bmodel%7D_%7Bk%2Cm%7D)%20-%20%5Coverline%7Bmodel%7D_%7Bk%2Cm%7D)#0>

where
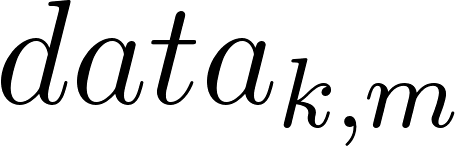
<https://www.codecogs.com/eqnedit.php?latex=data_%7Bk%2Cm%7D#0> and
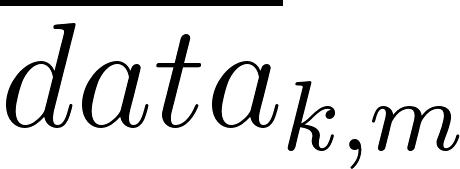
<https://www.codecogs.com/eqnedit.php?latex=%5Coverline%7Bdata%7D_%7Bk%2Cm%7D#0> are the numbers of term and preterm RSV hospitalisations respectively of children in age-group
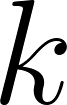
<https://www.codecogs.com/eqnedit.php?latex=k#0> and month
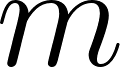
<https://www.codecogs.com/eqnedit.php?latex=m#0>, and
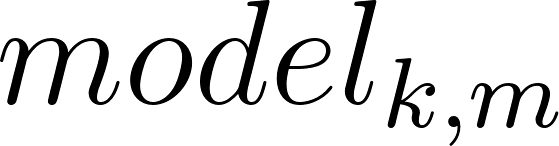
<https://www.codecogs.com/eqnedit.php?latex=model_%7Bk%2Cm%7D#0> and
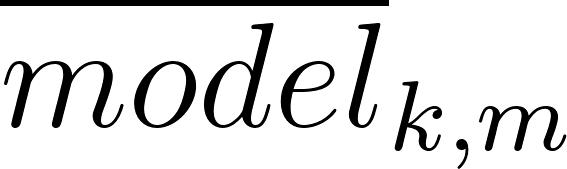
<https://www.codecogs.com/eqnedit.php?latex=%5Coverline%7Bmodel%7D_%7Bk%2Cm%7D#0> are the model predicted term and preterm RSV hospitalisation of children in age-group
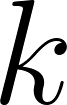
<https://www.codecogs.com/eqnedit.php?latex=k#0> and month
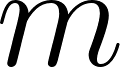
<https://www.codecogs.com/eqnedit.php?latex=m#0>for
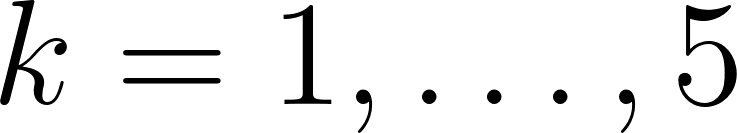
<https://www.codecogs.com/eqnedit.php?latex=k%3D1%2C%5Cldots%2C%205#0>. The subscript
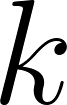
<https://www.codecogs.com/eqnedit.php?latex=k#0> for
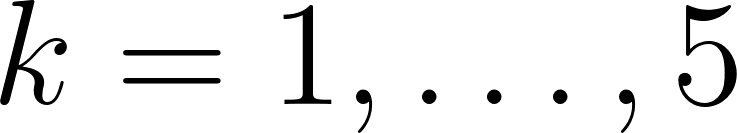
<https://www.codecogs.com/eqnedit.php?latex=k%3D1%2C%5Cldots%2C%205#0> denotes the five age-groups: 0-2 months, 3-5 months, 6-11 months, 12-23 months and 2-<5 years. For
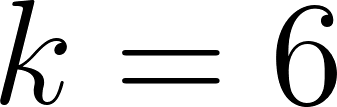
<https://www.codecogs.com/eqnedit.php?latex=k%3D6#0>,
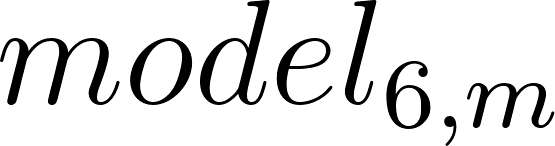
<https://www.codecogs.com/eqnedit.php?latex=model_%7B6%2Cm%7D#0> refers to the number of individuals of age 24-<25 months in month
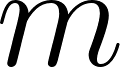
<https://www.codecogs.com/eqnedit.php?latex=m#0> that are in the naive susceptible compartment, i.e. the numbers of 2-year-olds that have never been infected, and
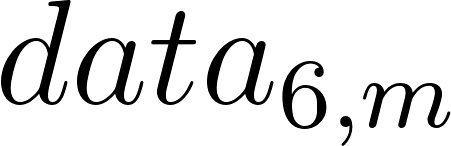
<https://www.codecogs.com/eqnedit.php?latex=data_%7B6%2Cm%7D#0> is 5% of the total number of individuals in this age group. Similar applies for the preterm equivalent variables.

#### Parameter estimates

The posterior samples of model parameters were generated from 8 independent chains of the Metropolis-Hastings sampler, each run for 5000 iterations after an initial, discarded ‘warm-up’ period of 1000 iterations per chain during which the sampler step size was also tuned. Convergence was assessed by visual assessment and diagnostic metrics, ensuring that the potential scale reduction factor for all parameters had values less than 1.1, and that there were at least 1000 effective samples across the 8 chains for each parameter (see Table S3). The estimated values of b_0_, b_1_ A, B and C resulting from fitting the model without the preterm stratification to the time series were then fixed in the stratified model before E, the preterm scaling factor, was estimated using the same fitting method as described with the RSV-hospitalisation time series separated into preterm and term births.

**Table S3:** Estimated maximum likelihood values for b_0_ the transmission coefficient, b_1_ the amplitude of the forcing function, and the age-to-risk function parameters: A, B, C and E. The Effective Sample Size (ESS) is the number of effectively independent draws from the posterior distribution that the Markov chain is equivalent to. The mean and 2.5, 25, 50, 75, and 97.5 percentile estimates are also provided.

| Parameter | ESS | Maximum likelihood estimate |
| --- | --- | --- |
| b_0_ | 2208.836 | 0.049356195 |
| b_1_ | 2552.217 | 0.143672161 |
| A | 2004.366 | 0.025995568 |
| B | 1712.354 | 0.185155023 |
| C | 1906.541 | 0.007098669 |
| E | 1104.725 | 2.548210491 |

|  | Mean | 2.5% | 25% | 50% | 75% | 97.5% |
| --- | --- | --- | --- | --- | --- | --- |
| b_0_ | 0.049361 | 0.049154 | 0.049290 | 0.049360 | 0.049431 | 0.049573 |
| b_1_ | 0.143532 | 0.138699 | 0.141857 | 0.143515 | 0.145189 | 0.148457 |
| A | 0.026315 | 0.023508 | 0.025263 | 0.026260 | 0.027299 | 0.029439 |
| B | 0.196762 | 0.148143 | 0.176523 | 0.194011 | 0.214410 | 0.259023 |
| C | 0.007260 | 0.006423 | 0.006970 | 0.007266 | 0.007551 | 0.008087 |
| E | 2.552091 | 2.349288 | 2.482160 | 2.547936 | 2.621745 | 2.757017 |

The model fitting calibrated the level of infection in the model such that at age 2 years, approximately 95% have experienced an RSV infection. As would be expected, the proportion of 2-year-olds that have been exposed at any point in a calendar year is dependent on birth month. Those that are born just before an RSV season, i.e. April-May, are less likely to have been exposed to RSV infection by age 2 than 2-year-olds born later in the year. This is likely due to those infants being born just before an RSV season having the highest level of natural maternal protection on entering their first peak RSV season. Table S4 shows the percentage of children at yearly ages up to 4 years old that have had at least one RSV infection, showing the range of this value over the year.

**Table S4:** The percentage of children at yearly ages up to 4 years old that have had at least one RSV infection, showing the minimum and maximum of this value over the year. The table also gives the percentage of mothers passing RSV antibodies to their infants as described and presented in Figure S7 and the maximum risk of hospitalisation as estimated through fitting the parameters of the age-to-risk function shown in Figure S6.

| **% at least one RSV infection** | | | | **% mothers passing RSV antibodies** | **Max. % of infections leading to hospitalisations** |
| --- | --- | --- | --- | --- | --- |
| 1 year | 2 years | 3 years | 4 years |  |  |
| 63.3 - 77.9  (May, Aug) | 92.4 - 96.2 (May, Sep) | 98.7 - 99.4 (Apr, Sep) | 99.8 - 99.9  (Apr, Sep) | 37.1 - 66.3  (Apr, Aug) | 3.31 (Term)  8.43 (Preterm) |

#### Model fit of hospitalisation predictions to observed data

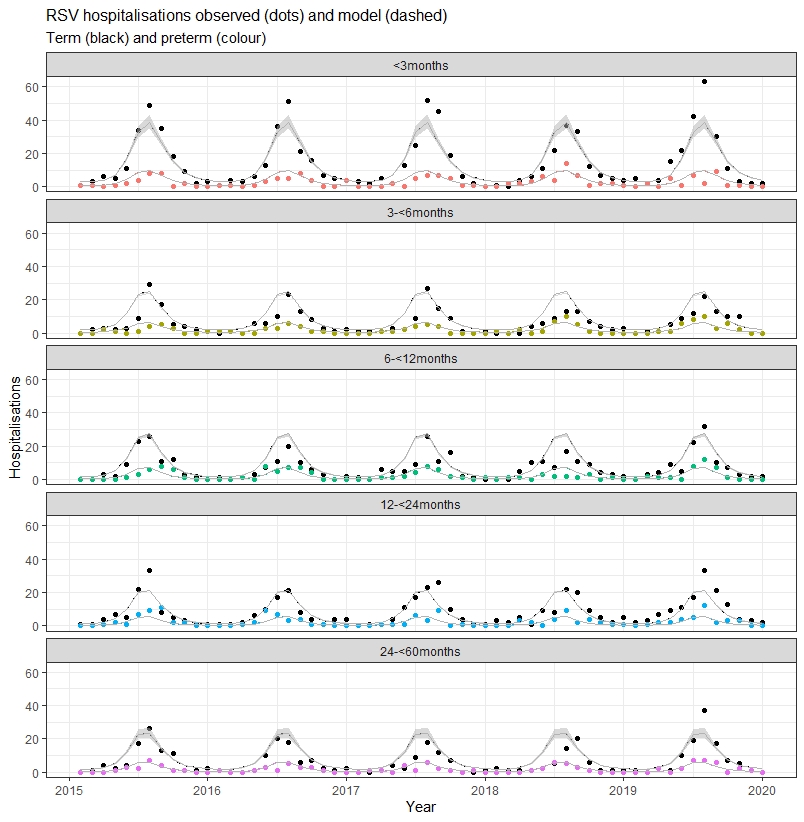

**Figure S3**: Comparison of the model estimated RSV-hospitalisations for the term (black) and preterm risk group (colour) to the observed time series of the five age groups used to fit the model. The observed hospitalisations are shown with dots, and the dashed line is the model output representing estimated hospitalisations. The grey band indicates the 95% confidence interval range of modelled output when the 2.5 and 97.5 percentile estimates for the fitted parameters are assumed (see Table S3).

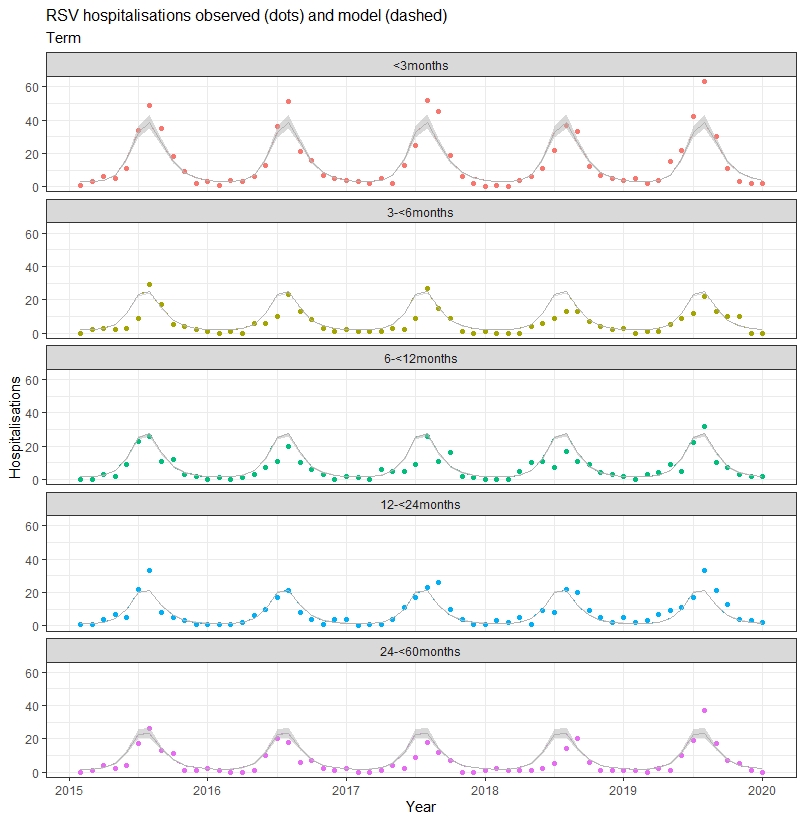

**Figure S4**: Comparison of the model estimated RSV-hospitalisations for children born at term to the observed time series of the five age groups used to fit the model. The observed hospitalisations are shown with dots, and the dashed line is the model output representing estimated hospitalisations. The grey band indicates the 95% confidence interval range of modelled output when the 2.5 and 97.5 percentile estimates for the fitted parameters are assumed (see Table S3).

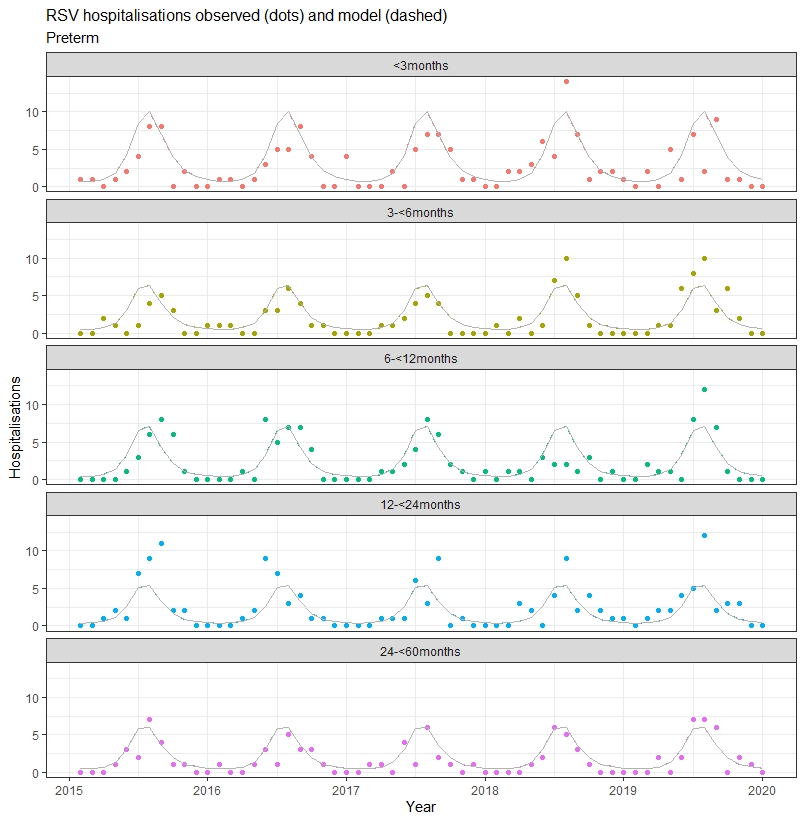

**Figure S5:** Comparison of the model estimated RSV-hospitalisations for the preterm risk group to the observed time series of the five age groups used to fit the model. The observed hospitalisations are shown with dots, and the dashed line is the model output representing estimated hospitalisations. The grey band indicates the 95% confidence interval range of modelled output when the 2.5 and 97.5 percentile estimates for the fitted parameters are assumed (see Table S3).

### Age-to-risk

**Figure S6:** The fitted age-to-risk exponential function relating age (in months) to risk of hospitalisation once infected by RSV. The red curves show risk for those infants born term and blue show the modified risk for preterm birth. Dashed lines relate to the decreased risk for second and subsequent infections while solid lines indicate the first infection. The grey band indicates the 95% confidence interval range of modelled output when the 2.5 and 97.5 percentile estimates for the fitted parameters are assumed (see Table S3).

### Modelled proportion of mothers passing on antibodies

Our fitted model captured the temporal dynamics of immunity to RSV in adults (Figure S7). In April, before the peak RSV months in temperate WA, the model predicted that about 37% of adults have some temporary immunity to RSV due to recent infection. After the RSV peak, about 66% of 15-45-year-olds have been recently infected. This implies that adults in this age range are infected with RSV approximately every 1 in 3 RSV seasons. We assume the minimum value as an estimate of the proportion of mothers passing on RSV antibodies to their newborns (see Figure S8 for sensitivity analysis assuming maximum value), and with the exponential decay of maternal antibody protection, scale the susceptibility to RSV infection of infants under 12 months accordingly (see Table S2).

**Figure S7**: The modelled proportion, over a calendar year, of adults between the ages of 15 to 45 years that were not in a susceptible compartment and hence have been recently exposed to RSV infection. This is used as an estimate of the proportion of mothers passing on RSV antibodies to newborns.

### Immunisation protection profiles

#### mAb

To simulate the protection profile of mAb (nirsevimab) in the model, we assume an Erlang-3 distribution, implemented by “chaining” three exponentially- distributed protected compartments [[20]](https://paperpile.com/c/Y68o4Y/IEcO) (see Figure 1). Those waning from the first protected compartment move into a second and then third protected compartment, if they are not infected, before returning to an unprotected susceptible population. Chaining three monoclonal protected compartments means that, on average, the duration of protection of those receiving nirsevimab more closely follows the Erlang-3 distribution with the same mean protection period of 150 days, but with a more sustained level of protection in the earlier period and more marked drop-off of protection after 150 days as compared to the exponential distribution with the same mean (see Figure S8). An average of 78% are protected over the 150 days immediately following immunisation, compared to 63% for the exponential distribution corresponding to one protected compartment, and the proportion of those still protected drops to ~5% at 300 days rather than ~14% (Figure S8). This average of 78% protected over the first 150 days aligns well with the assumed efficacy [[18]](https://paperpile.com/c/Y68o4Y/H2PRX), and hence we assume that if those in a nirsevimab-protected susceptible compartment are infected, there is no risk of hospitalisation. Those in protected compartments that are infected, follow the infection path in the model with the assumption that temporary protection to further infection then comes from immunity due to infection rather than from nirsevimab (Figure 1).

**Figure S8**: Increasing the number of mAb protected compartments from k = 1 to maximum of k = 10 and finding the resulting Erlang distribution, assuming a mean durability of 150 days post immunisation. Dashed lines give the average proportion of vaccinees protected over the first 150 days for each of the distributions.

#### Maternal vaccine

To simulate the protection of the maternal vaccine (Abrysvo) in the model, we explored the number of compartments and average durability assumptions that best matched clinical trial efficacy estimates [[21]](https://paperpile.com/c/Y68o4Y/J2aaK) associated with RSV-hospitalisations. The efficacy estimates associated with RSV-hospitalisations 90, 120, 180 and 360 days after birth are 68%, 60%, 57% and 33% respectively, and are indicated by the black dashed line in Figure S9. An exponential decay profile, which corresponds to one protection compartment in the model for the maternal vaccine (Figure 1), most closely matched the decay profile of the efficacy estimates. We then explored three exponential distributions with average durability of 100, 120 and 150 days (red, green and blue curves in Figure S9). The green point estimate and the associated lower and upper values of the interval bar correspond to the average proportion of vaccinees protected over the first 90, 120, 180 and 360 days for each of these distributions. The exponential waning profile with average durability of 120 days (associated with the green point estimates), most closely matched the efficacy estimates and hence was used in the model.

**Figure S9** : Plot illustrating the process of “fitting” a waning protection profile to the clinical trial estimates of the efficacy over 90, 120, 180 and 360 days associated with RSV-hospitalisation of maternal vaccine [[21]](https://paperpile.com/c/Y68o4Y/J2aaK). These estimates are provided in black alongside the dashed lines. The red, green and blue curves relate to exponential distributions assuming a mean durability of 100-, 120- and 150-days post birth. The green dots are the average proportion of vaccinees protected over 90, 120, 180 and 360 days for the exponential distribution with mean 120 days, the lower limit of the bars correspond to the same values for the exponential distribution with mean of 100 days, and the upper limit corresponds to the values associated with the exponential distribution with mean of 150 days.

Comparison

**Figure S10**: Comparison of mAb and maternal vaccine waning protection profiles

### Scenario output

**Table S5**: Hospitalisations averted, and proportion of hospitalisations averted yearly for under 3-month, 3-<6 months, 6-<12 months and under 24-month children, the number of yearly nirsevimab and Abrysvo doses, and the number needed to immunise (NNI) to prevent one RSV-hospitalisation, for all modelled scenarios.

| **Scenario** | | | **Coverage** | **Hospitalisations averted** | | | | **Prop. of hospitalisations averted** | | | | **Number of doses** | | | **NNI** |
| --- | --- | --- | --- | --- | --- | --- | --- | --- | --- | --- | --- | --- | --- | --- | --- |
|  |  |  |  | <3m | 3-<6m | 6-<12m | <24m | <3m | 3-<6m | 6-<12m | <24m | mAb | mat vac | total |  |
| **mAb** | 1(a) | **Year-round at birth** | 50% | 88.84 | 33.01 | 12.69 | 135.64 | 0.43 | 0.25 | 0.09 | 0.23 | 15381 | 0 | 15381 | 113 |
|  | 1(b) | **Year-round at birth** | 70% | 124.38 | 46.21 | 17.77 | 189.89 | 0.60 | 0.35 | 0.13 | 0.33 | 21533 | 0 | 21533 | 113 |
|  | 1(c) | **Year-round at birth** | 90% | 159.92 | 59.42 | 22.85 | 244.15 | 0.78 | 0.46 | 0.17 | 0.42 | 27685 | 0 | 27685 | 113 |
|  | 2(a) | **Seasonal at birth** | 50% | 67.91 | 15.79 | 3.62 | 87.84 | 0.33 | 0.12 | 0.03 | 0.15 | 7690 | 0 | 7690 | 88 |
|  | 2(b) | **Seasonal at birth** | 70% | 95.07 | 22.11 | 5.07 | 122.98 | 0.46 | 0.17 | 0.04 | 0.21 | 10766 | 0 | 10766 | 88 |
|  | 2(c) | **Seasonal at birth** | 90% | 122.23 | 28.43 | 6.52 | 158.12 | 0.59 | 0.22 | 0.05 | 0.27 | 13843 | 0 | 13843 | 88 |
|  | 3 | **Seasonal at birth with catchup + 2nd season high-risk** (2024 WA program) | 79% - birth, 65% - catchup,  30% - high-risk | 115.60 | 51.52 | 33.73 | 206.05 | 0.56 | 0.39 | 0.24 | 0.35 | 19389 | 0 | 19389 | 94 |
| **mat vac** | 4(a) | **Year-round** | 50% | 70.03 | 22.82 | 10.28 | 104.72 | 0.34 | 0.17 | 0.07 | 0.18 | 0 | 15381 | 15381 | 147 |
|  | 4(b) | **Year-round** | 70% | 98.04 | 31.95 | 14.40 | 146.61 | 0.48 | 0.24 | 0.10 | 0.25 | 0 | 21533 | 21533 | 147 |
|  | 4(c) | **Year-round** | 90% | 126.05 | 41.08 | 18.51 | 188.50 | 0.61 | 0.31 | 0.13 | 0.32 | 0 | 27685 | 27685 | 147 |
|  | 5(a) | **Seasonal** | 50% | 54.44 | 10.92 | 3.19 | 69.41 | 0.26 | 0.08 | 0.02 | 0.12 | 0 | 7690 | 7690 | 111 |
|  | 5(b) | **Seasonal** | 70% | 76.21 | 15.29 | 4.47 | 97.18 | 0.37 | 0.12 | 0.03 | 0.17 | 0 | 10766 | 10766 | 111 |
|  | 5(c) | **Seasonal** | 90% | 97.99 | 19.66 | 5.74 | 124.94 | 0.48 | 0.15 | 0.04 | 0.21 | 0 | 13843 | 13843 | 111 |
| **hybrid** | 6(a) | **Year-round mat vac + seasonal mAb at birth high-risk** | mat vac - 50%  mAb - 100% (high-risk) | 86.73 | 27.06 | 11.11 | 126.53 | 0.42 | 0.21 | 0.08 | 0.22 | 1415 | 14673 | 16088 | 127 |
|  | 6(b) | **Year-round mat vac + seasonal mAb at birth high-risk** | mat vac - 70%  mAb - 100% (high-risk) | 110.27 | 35.30 | 14.97 | 162.72 | 0.53 | 0.27 | 0.11 | 0.28 | 1415 | 20542 | 21957 | 135 |
|  | 6(c) | **Year-round mat vac + seasonal mAb at birth high-risk** | mat vac - 90%  mAb - 100% (high-risk) | 133.81 | 43.53 | 18.82 | 198.91 | 0.65 | 0.33 | 0.14 | 0.34 | 1415 | 26412 | 27827 | 140 |
|  | 7(a) | **Year-round mat vac OR seasonal mAb at birth** | mat vac - 30%  mAb - 50% | 109.92 | 29.49 | 9.79 | 150.68 | 0.53 | 0.23 | 0.07 | 0.26 | 7690 | 9228 | 16919 | 112 |
|  | 7(b) | **Year-round mat vac OR seasonal mAb at birth** | mat vac - 45%  mAb - 35% | 110.56 | 31.60 | 11.79 | 155.74 | 0.54 | 0.24 | 0.09 | 0.27 | 5383 | 13843 | 19226 | 123 |
|  | 7(c) | **Year-round mat vac OR seasonal mAb at birth** | mat vac - 60%  mAb - 20% | 111.19 | 33.71 | 13.79 | 160.81 | 0.54 | 0.26 | 0.10 | 0.28 | 3076 | 18457 | 21533 | 134 |
|  | 8(a) | **Year-round mat vac OR seasonal mAb at birth + 2nd season mAb high-risk** | mat vac - 30%  mAb - 50% (at birth), 30% (high-risk) | 109.92 | 29.49 | 11.50 | 155.44 | 0.53 | 0.23 | 0.08 | 0.27 | 8229 | 9228 | 17458 | 112 |
|  | 8(b) | **Year-round mat vac OR seasonal mAb at birth + 2nd season mAb high-risk** | mat vac - 45%  mAb - 35%  (at birth), 30% (high-risk) | 110.56 | 31.60 | 13.47 | 160.46 | 0.54 | 0.24 | 0.10 | 0.28 | 5919 | 13843 | 19761 | 123 |
|  | 8(c) | **Year-round mat vac OR seasonal mAb at birth + 2nd season mAb high-risk** | mat vac - 60%  mAb - 20%  (at birth), 30% (high-risk) | 111.19 | 33.71 | 15.43 | 165.47 | 0.54 | 0.26 | 0.11 | 0.28 | 3609 | 18457 | 22065 | 133 |
|  | 9(a) | **Year-round mat vac OR seasonal mAb at birth + catchup + 2nd season mAb high-risk** (2025 WA program) | mat vac - 30%  mAb - 50% (at birth), 70% catchup, 30% (high-risk) | 117.24 | 53.43 | 36.24 | 212.67 | 0.57 | 0.41 | 0.26 | 0.36 | 14433 | 9228 | 23661 | 111 |
|  | 9(b) | **Year-round mat vac OR seasonal mAb at birth + catchup + 2nd season mAb high-risk** (2025 WA program) | mat vac - 45%  mAb - 35%  (at birth), 70% catchup, 30% (high-risk) | 117.11 | 53.48 | 36.76 | 213.35 | 0.57 | 0.41 | 0.27 | 0.37 | 11690 | 13843 | 25532 | 120 |
|  | 9(c) | **Year-round mat vac OR seasonal mAb at birth + catchup + 2nd season mAb high-risk** (2025 WA program) | mat vac - 60%  mAb - 20%  (at birth), 70% catchup, 30% (high-risk) | 116.98 | 53.53 | 37.28 | 214.03 | 0.57 | 0.41 | 0.27 | 0.37 | 8947 | 18457 | 27403 | 128 |
